# The contribution of evolutionary game theory to understanding and treating cancer

**DOI:** 10.1101/2020.12.02.20241703

**Authors:** Benjamin Wölfl, Hedy te Rietmole, Monica Salvioli, Artem Kaznatcheev, Frank Thuijsman, Joel S. Brown, Boudewijn Burgering, Kateřina Staňková

## Abstract

Evolutionary game theory mathematically conceptualizes and analyzes biological interactions where one’s fitness not only depends on one’s own traits, but also on the traits of others. Typically, the individuals are not overtly rational and do not select, but rather inherit their traits. Cancer can be framed as such an evolutionary game, as it is composed of cells of heterogeneous types undergoing frequency-dependent selection. In this article, we first summarize existing works where evolutionary game theory has been employed in modeling cancer and improving its treatment. Some of these game-theoretic models suggest how one could anticipate and steer cancer’s eco-evolutionary dynamics into states more desirable for the patient via evolutionary therapies. Such therapies offer great promise for increasing patient survival and decreasing drug toxicity, as demonstrated by some recent studies and clinical trials. We discuss clinical relevance of the existing game-theoretic models of cancer and its treatment, and opportunities for future applications. Moreover, we discuss the developments in cancer biology that are needed to better utilize the full potential of game-theoretic models. Ultimately, we demonstrate that viewing tumors with an evolutionary game theory approach has medically useful implications that can inform and create a lockstep between empirical findings and mathematical modeling. We suggest that cancer progression is an evolutionary game and needs to be viewed as such.

## 1. Introduction & Motivation

Cancer is a disease of unregulated proliferation, caused by abnormal function of genes responsible for regulating cell division. The genesis of cancer has strong ties to the human life history [6, 67, 90, 91, 174] and its progression is driven by natural selection, characterized by cancer cells exhibiting the following three conditions [55]:

1. The presence of heritable variation: Heritable traits vary among different cancer cells, ultimately as a result of genetic mutation.
2. A struggle for existence: There are limits to growth due to competition for limited space and resources.
3. The influence of heritable variation on the struggle for existence: Generally, the likelihood of cell survival depends on its own traits, and the traits of the others. Cells with traits that confer higher chances of survival and proliferation will in time increase in frequency (frequency-dependent selection) [90, 91].

This Darwinian view of cancer is in line with the premises of evolutionary game theory (EGT), which assumes that evolution tests heritable traits in an ongoing competition for survival [36, 97, 123, 124]. EGT is a branch of mathematics that has helped to conceptualize and understand the behavior of real-world biological systems, including several counter-intuitive biological phenomena [88, 89, 124, 157, 177, 195] and is being increasingly recognized as an important tool for mathematical oncologists [18, 149].

EGT deals with situations where organisms using different strategies and/or possessing different traits interact with each other. Unlike in classical game theory [135, 184, 185], these organisms do not need to be overtly rational, i.e., their strategies (often referred to as “types”) are inherited rather than rationally chosen [35, 97] (although a rational population-level interpretation of the dynamics is also possible [105]). Some strategies might confer higher fitness and the individuals using these strategies will in a long run dominate the population. Thus, if we see cancer as a Darwinian process, it can be described as an evolutionary game, where cancer cells are the players, their heritable traits correspond to the strategies, and the payoffs are represented in terms of survival and proliferation (fitness) [36, 125]. This is a dynamic game, as one can analyze how frequencies of different strategies and numbers of individuals corresponding to these different strategies change in time. We refer to those changes as evolutionary and ecological dynamics, respectively. Both together are called eco-evolutionary dynamics.

Compared to other fields of applied mathematics, EGT of cancer is a relatively novel field, just few decades old [119, 176]. Tomlinson (1997) was first to explicitly frame cancer as an evolutionary game and since then at least 60 publications on cancer have called their research game-theoretic. This body of literature has grown into diverse and different groupings. Given that cancer is an evolutionary process, it has been suggested that also cancer treatment could benefit from insights from evolutionary theory, giving rise to Evolutionary or Darwinian medicine [74, 76, 80]. The increasing interest in this field is reflected in the recent update of medical curricula to include evolutionary reasoning [136]. Clearly, application of EGT can only improve cancer treatment if there is something gained from these evolutionary insights. Standard of Care (SoC) in treating cancer typically applies therapy at the Maximum Tolerable Dose (MTD), to remove tumor cells as fast as possible. While for some aggressive cancers, such as, for example, advanced Non-Small Cell Lung Cancer, no better treatment than MTD has been found so far [20, 9, 19], it has been also observed that unless the patient cures, the MTD strategy promotes evolution of treatment-induced resistance which often leads to treatment failure [73, 200, 159].

The fact that even personalized therapies tailored to the cancer’s genetic signature, and to the individual’s genetic disposition fail can be attributed to the extensive adaptive potential of the human genome. As MTD can only eradicate therapy-sensitive tumor cells, it may lead to the benefit of therapy-resistant cells [79, 142]. Subsequently, growth-limiting constraints due to competition may temporarily vanish and increase the *per capita* growth rate of the resistant types (competitive release [47, 65, 159, 198]). In turn, some experiments show that when treatment is stalled (drug holiday), resistant types are typically at a disadvantage (cost of resistance [163]; although this is not universal [109]). This evidence suggests that MTD might be evolutionary unwise if it promotes treatment-induced resistance in cancer cells. Additionally, there is evidence for selection for evolvability in tumor cells, e.g., hyper-mutators [41]. Recent works showed that a game-theoretic approach may help to provide an alternative to MTD, based on anticipating and steering the cancer eco-evolutionary dynamics in response to the treatment [85, 160].

The aims of this paper are the following: (i) to discuss the achievements of the existing works on game theory of cancer and (ii) to show the future potential of game theory to understand cancer mechanisms, inspire novel research, and design better treatment protocols.

In the remainder of this paper, we will firstly introduce models where the interaction among cells is explicitly framed as an evolutionary game, with either no or fixed treatment (Section 2). Subsequently, we will review cancer models where the physician as a rational player optimizing their own objective(s) enters the evolutionary game (Section 3). Thirdly, we will focus on the clinical aspects of EGT therapy models (Section 4). We will close this paper with a discussion on limitations and future steps in game theory of cancer and its treatment (Section 5).

### 1.1. Mathematical background

Cancer is a Darwinian disease, in which cancer cells play an evolutionary game between each other within the dynamic environment of the tumor that also includes diverse normal cells (stroma) [21, 81, 176]. The cells may have different types, varying in their (possibly evolving) level of resistance to a particular treatment or treatment combination [74, 81, 120, 159]. Here we do not specify whether these types are just phenotypically or also genetically different - that is why we confine ourselves to the term “types”, as opposed to “clones” used in some literature. For some cancers, such as metastatic Castrate-Resistant Prostate Cancer (mCRPC) and Estrogen Receptor Positive (ER+) breast cancer, cancer types differing in their resistance levels with respect to a particular treatment have been identified both *in vitro* and *in vivo* [64, 70, 80, 201]. For less researched cancers, such types have not been established yet and it may be that resistance varies per cancer cell, as a resistance trait evolves in response to treatment.

Therefore, in the most general game-theoretic model of cancer, the resistance of a particular cancer cell type to a particular treatment is a continuous evolving heritable trait. Then, individual cancer cells are identified by their value of this trait, which is subject to natural selection. Here we will adopt the *Darwinian dynamics* approach to describe such a situation, expanding the original model of Vincent and Brown (2005) into more dimensions [180].

A vector **x**(*t*) = (*x*_1_(*t*), …, *x*_*n*_(*t*))^*T*^ defines population densities (population size per space) of cancer cells of types *𝒯* = {1, …, *n*} at time *t*. The fitness of cancer cells of type *i* ∈ *𝒯* may depend on the densities and traits of all cancer cell types. Consequently, the ecological dynamics of cancer cells of type *i* are given by

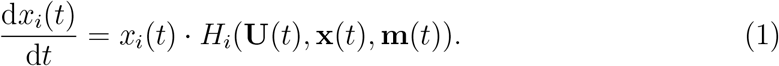

Here, **U**(*t*) = (*u*_*ij*_(*t*)) is a resistance matrix, where *u*_*ij*_(*t*) ∈ [0, 1] indicates the resistance level of cancer cells of type *i*, in response to treatment *j*. Moreover, **m**(*t*) = (*m*_1_(*t*), …, *m*_*p*_(*t*))^*T*^ is the vector of doses for each therapy option from the treatment set Θ. Without loss of generality, we can assume that *m*_*j*_(*t*) ∈ [0, 1] for all *j* ∈ Θ, where *m*_*j*_(*t*) = 1 and *m*_*j*_(*t*) = 0 correspond to the MTD and no dose of treatment *j* at time *t*, respectively. In this formulation, we see that the *per capita* growth rate *H*_*i*_(**U**(*t*), **x**(*t*), **m**(*t*)) of type *i* may give rise to density and frequency-dependent dynamics, as it depends on **x**(*t*) explicitly.

Vincent and Brown (2005) developed the concept of a fitness generating function as a way to describe the fitness of many species (or types) by making use of a single mathematical expression [180]. A function *G*(*v*, **U**(*t*), **x**(*t*), **m**(*t*)) is said to be a *fitness generating function* (G-function) of the population dynamics (1) if

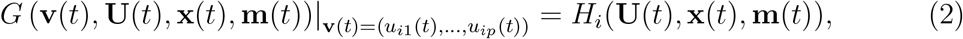

where **v** is a virtual vector variable. Replacing *v*_*j*_ in the G-function with *u*_*ij*_ for each *j* ∈ Θ yields the fitness of an individual cell of type *i* in a population defined by the same G-function. Using the G-function, we can rewrite equation (1) as:

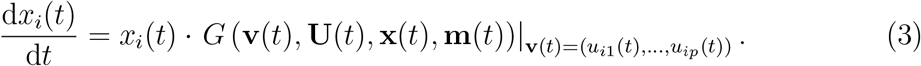

Cancer types with a higher *per capita* growth rate will persist in the population. Therefore, the dynamics of the evolution of resistance *u*_*ij*_ of the cancer cell of type *i* in response to a treatment *j* (evolutionary dynamics) is given as follows:

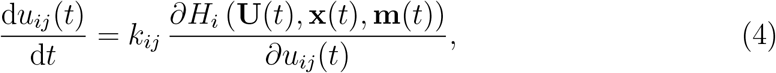

which can be rewritten using the G-function as:

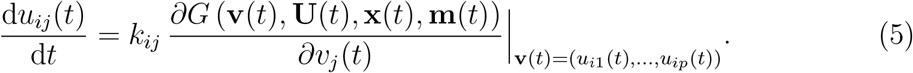

Here, *k*_*ij*_ is a speed parameter, which is a measure of heritability and additive genetic variance, in line with Fisher’s fundamental theorem of natural selection [66]. This speed parameter may be influenced by many other factors, like mutation rates, population size, population structure and the underlying genetics of inheritance. For example, in *adaptive dynamics, k*_*ij*_ is linearly increasing with population size and stochastic with respect to other parameters (canonical equation of adaptive dynamics [58, 83, 92, 126]). For the sake of simplicity, when modeling (5), it is often assumed that *k*_*ij*_ is the same constant for all *i* and *j*, while one could easily imagine that *k*_*ij*_ varies in time and may be a (likely nonlinear) function of *x*_*i*_(*t*). In the remainder of this paper, we will not write out the time-dependence explicitly, thus we shall use **U, x** and **m** instead of **U**(*t*), **x**(*t*) and **m**(*t*), respectively. Equations (3) and (5) constitute the Darwinian dynamics, describing the ecological and evolutionary dynamics of cancer cells, respectively.

If the ecological dynamics (3) converge to a stable equilibrium **x**\* ≥ 0, we call **x**\* an *ecological equilibrium*. Each combination of resistance and treatment strategies (**U, m**) may have an associated vector of stable population sizes **x**\*, with 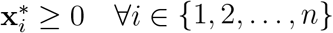. A generic **U** may have one or more values of **x**\*, or no equilibrium associated with it, depending on the G-function. Moreover, even if we assume that the ecological equilibrium exists for any choice of **U** and **m**, it may be that only a subset of possible values of **U** and **m** will correspond to positive equilibrium population sizes. Depending upon the model, its parameters and the strategies **U** and **m**, there will likely be an upper limit to the number of types that can co-exist at positive population sizes [87].

Solved together for **m** fixed at particular values, equations (3) and (5) often determine an equilibrium solution for **x**(**m**) and **U**(**m**), which we will denote by **x**\*(**m**) and **U**\*(**m**), respectively. The non-zero equilibrium values of 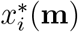 and their associated strategies 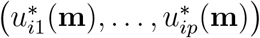 form a ‘coalition’ of strategies. If, for a particular choice of **m**, these strategies resist invasion by other mutant strategies, they are called Evolutionarily Stable Strategies (ESSs) with respect to treatment **m** [97]. A necessary condition for an ESS is that the G-function maximizes *G* with respect to **v** at the corresponding **x**\*. Further stability properties of the ESS exist (e.g., convergence stability or neighborhood invasion stability [10, 11]).

In Section 2, we will consider existing models of cancer *without treatment* and those that consider a *predefined fixed treatment*. Such treatments may administer a constant dose treatment **m**(*t*) or, for example, pause treatment when the total tumor population is below a certain predefined threshold, and re-administering it again once the population of tumor cells recover to its initial size. In Section 3, we will consider situation where the physician enters the ‘game against cancer’ as a rational player, i.e. a player optimizing certain objective(s) with respect to their treatment strategies.

## 2. Game theory of cancer without treatment or with a predefined treatment regimen

In the literature on EGT models of cancer with no or predefined treatment **m**, the authors either focus on finding the ESS resistance strategy **U**\* at the ecological equilibrium **x**\*, or they analyze transient dynamics towards (**x**\*, **U**\*) for particular (predefined) choices of **m**, to see what choices of **m** are better than others in terms of some prespecified metrics, such as progression-free or overall survival.

Firstly, we will present a paper that describes a model in the form of equations (3) and (5), followed by models that somewhat simplify the two equations by using a fitness and a competition matrix, respectively, and spatial models.

### 2.1. Models with eco-evolutionary dynamics described by equations (3) and (5)

In their commentary on treating of pediatric sarcoma, Reed et al (2020) introduce a model using pediatric sarcoma’s G-function

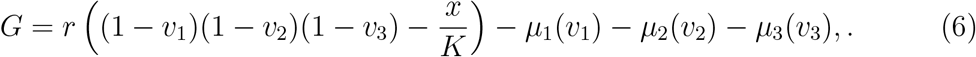

Here *v*_*i*_ denotes the treatment-induced resistance to treatment *i, r* is the intrinsic growth rate of the tumor cells, and 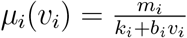 is the treatment-induced death rate for treatment *i*. In *µ*_*i*_(*v*_*i*_), *m*_*i*_ is the base treatment-induced death rate of the tumor cells, *k*_*i*_ denotes innate resistance, and *b*_*i*_ gives the benefit gained by accumulating resistance towards drug *i*. Reed et al adopt the framework given by (5) and (3) with a G-function defined by (6) to analyze possible strategies to combat the pediatric sarcoma, motivated by the theory of extinction from ecology, recently discussed in the oncology literature as well [82, 75].

Motivated by numerical simulations on different treatment regimens, the authors suggest that when a cure for pediatric sarcoma is an achievable outcome, the first strike (standard of care) therapy should be either augmented, or closely followed with diverse second strike therapies. They hypothesize that application of the “first-strike” and “second-strike” therapies may improve the standard of care, which relies on fixed combination of chemotherapies.

In contrast, when it is believed that a cure is unachievable, they propose the adaptive therapy protocol proposed by Zhang et al (2017) for metastatic castrate-resistant prostate cancer, which we will discuss in Section 2.5 [200].

### 2.2. Replicator dynamics with fitness matrix

The simplest and often very intuitive game-theoretic cancer models are those where the fitness of cancer cells is given by a fitness matrix. These models typically assume that the cancer cells engage in pairwise interactions and as a result of these interactions, the cells may reproduce, generating offspring of the same type as the parental cell (although other interpretations are also possible [105, 109]).

Let us denote by *a*_*ij*_ the expected number of offspring generated by a cancer cell of type *i* interacting with a cell of type *j*. Alternatively, if *a*_*ij*_ ∈ [0, 1], it can define the probability of a cell of type *i* producing an offspring of its own type when interacting with a cell of type *j*. If we have *n* types of cancer cells and we know *a*_*ij*_ for all *i, j* ∈ {1, …, *n*}, we can construct an *n* × *n* fitness matrix **A** = (*a*_*ij*_). The population (ecological) dynamics of cells of different types as proportions, **q**, instead of densities, **x**, are commonly described by *replicator dynamics* [25, 26, 105, 109, 111, 167, 168, 188] where 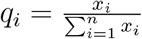 is defined by

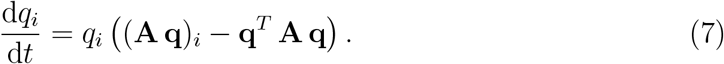

Here, the *per capita* growth rate of cancer cells of type *i* is given by their expected payoff (fitness) (**A q**)_*i*_ minus the mean fitness of the entire population **q**^*T*^ **A q**. This fitness is frequency-dependent [46, 108] and captures non-cell-autonomous effects that are central to the ecology of cancer [121, 109, 62].

The replicator dynamics represent a special case of (3) and (5) as it considers population (ecological) dynamics only in terms of proportions and does not consider the evolutionary dynamics of different types of cancer cells. The latter point implies that it fits within the framework set by equation (3) with the G-function defined by (**A q**)_*i*_ − **q**^*T*^ **A q**, where trait **U** simply does not evolve. The dynamics of frequencies *q*_*i*_ with *i* = 1, …, *n* are restricted to the *n*-dimensional simplex, i.e., Σ*i q*_*i*_ = 1 which means that the 0 ≤ *q*_*i*_ ≤ 1 are proportions. Accordingly, an average fitness above 1 cannot yield an increase in total density and, therefore, the effects of density-dependent selection are not modelled. Instead, solely frequency-dependent selection is considered.

As shown by Zeeman (1980), any ESS of matrix **A** is an attractor (stable equilibrium) of the replicator dynamics (7) [197]. If such an ESS in tumors exists, reaching it using available therapies could provide a means for achieving *long-term stabilization* of tumors and a significant increase in progression-free and overall survival [109, 188, 53]. However, it is important to be aware of the timescales involved. In the most general case, even on very long timescales fixed points like ESS might not be reached [46, 107], for example due to the evolutionary constraints of population size [84].

One of the first models that defines the competitive interactions of cancer cells via a fitness matrix following Equation (7), was called ‘Go-vs-Grow game’ and introduced by Basanta et al. [24] and promptly extended to include glycolysis [26]. Here, the interaction between three cell cancer types of invasive (Go), autonomous growth (Grow) and glycolytic (GLY) types was introduced and it was analyzed for how different parameters in the fitness matrix **A** influence the game characteristics and ESSs [26]. The main outcomes of this analysis are that an invasive cancer type is more likely to evolve after occurrence of the glycolytic type, and that the therapies increasing the fitness cost of switching to anaerobic glycolysis might decrease the probability of the emergence of more invasive cancer type. The follow-up work includes stromal cells interacting with different types of cancer cells and their role in promoting cancer invasiveness [25].

Dingli et al. (2009) showed that targeting the interactions between the tumor and the stromal cells, so that the latter outcompete the former ones, can be a more promising approach, compared to targeting the cancer cells directly [61]. Other examples of cancer games with the fitness defined by a matrix are the cooperative ones, following the paper of Axelrod et al. (2006) summarizing evidence of cooperation among cancer cells [21].

### 2.3. Estimating parameters of the fitness matrix

Although the above works modelled the interactions among cancer cells of different types and their environment as a fitness matrix, the parameters of these matrices were not directly measured or *in vivo* or *in vitro*. To remedy this, Kaznatcheev et al. (2019) introduced a technique to directly estimate parameters of the fitness matrix of replicator dynamics [109] from data measured *in vitro*. They studied interactions of different cancer cell types in co-cultures of non-small cell lung cancer (NSCLC) cells [109]. The cancer cell types included those sensitive (parental) and resistant to the anaplastic lymphoma kinase inhibitor alectinib. With two cell types, the replicator dynamics describing the change in frequencies of the parental, *p*, and resistant, 1 − *p*, cancer cell types in the population, becomes:

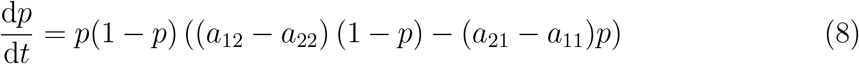

with a fitness matrix **A** = (*a*_*ij*_).

Since certain changes in the coefficients of the matrix **A** imply changes of mathematical properties of Equation (8), such as existence of ESS, four different dynamic regimes are possible. Kaznatcheev et al. (2019) estimated the entries of the fitness matrix **A** from the growth data of a series of specifically-designed *in vitro* experiments across four different environmental conditions corresponding to the presence or absence of targeted therapy and the presence and absence of cancer-associated fibroblasts. They show that the games played by the population *in vitro* produce two qualitative different dynamics regimes, i.e., that the dynamics (8) qualitatively switch the type of game being played by the population *in vitro* from a game they term a ‘Deadlock game’ to a game they term a ‘Leader game’, based on the presence of absence of drug and/or fibroblasts.

While therapy optimization was not the goal of this study (in fact, therapy eventually failed for all considered cases), Kaznatcheev et al.’s provide the game assay as a method to estimate the enties in the fitness matrix from *in vitro* data. This allows to anticipate treatment-induced eco-evolutionary responses of cancer cells even before the treatment is applied in order to steer eco-evolutionary dynamics of cancer cells during the course of the treatment [109, 159]. Subsequent work focused on quantifying competitive release in NSCLC [65] and extended the original game to a game with 3 types of cancer cells [32]. A similar method was used to observe host-parasite-like interactions between cancer cell types due to paracrine behaviors [138].

### 2.4. Replicator dynamics with non-linear fitness functions

Although the system studied by Kaznatcheev et al. (2019) is well estimated by replicator dynamics with fitness given by the linear function (**Aq**)_*i*_, their method can also be used to estimate parameters in non-linear fitness functions, i.e. a generalization of of (7)

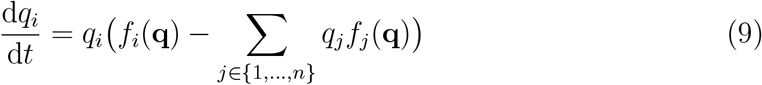

where the fitness functions *f*_*i*_ are not necessarily linear.

This case of non-linear fitness functions has generated extensive theoretical work in the case of public goods where cells can be producers (cooperators) or free-ride on shared resources produced by the others (defectors) [16]. The most relevant cases for cancer are the production of growth factors like VEGF [12, 154], the production of hostile environments like acidity due to the Warburg effect [43, 13, 14], or the coupling of both [111]. Archetti et al (2015) empirically estimated parameters of this nonlinear public good game for neuroendocrine pancreatic cancer cells that produce insulin-like growth factor II which supports proliferation and evasion of apoptosis, [17].

For the situation when the public good of VEGF production is coupled with the public good of of tumour acidity, Kaznatcheev et al (2017) targeting the most common cancer cell type through MTD may lead to a worse long-term outcome for the patient than targeting less common types [111].

### 2.5. Lotka-Volterra models

As the replicator dynamics (9) include frequency dynamics only and cannot capture the changes of cancer cell population, alternative modeling frameworks have been adopted to capture both ecological and evolutionary dynamics of cancer cells. These include expanding replicator dynamics by additional population dynamics elements, such as through fictious free-space strategies [104, 187], but also different models capturing population dynamics directly [119, 180]. A relatively large body of literature models interactions between cancer cells of different types and/or interactions between cancer cells and the environment through the Lotka-Volterra competition equations and their extensions [29, 52, 72, 200]. The LV equations were proposed separately by Lotka and Volterra to describe competition in one set of models and predator-prey dynamics in another one [116, 183]. Here we restrict ourselves to the competition models.

While initially the LV dynamics described interactions between two species only, they can also be expanded to model interactions of cancer cells of *n* types. Moreover, it is possible to convert the replicator dynamics for *n* types into the LV model with *n* − 1 types and vice versa, by converting the fitness matrix **A** into the *competition matrix* of the LV model and maintaining the same stable equilbria (attractors). The proof of this ESS equivalence can be found e.g. in [97] and [35]. The attractors of the LV dynamics correspond to the attractors of the replicator dynamics (7) and may correspond to the ESSs of the matrix **A**, as discussed before. For instance, The ESSs of You et al’s replicator dynamics model of mCRPC in [193] are the same as the ESSs of the LV model in [200]. The LV model describes ecological dynamics (3), while the evolutionary dynamics are trivial as the resistance trait does not evolve and therefore corresponds to (5) with the right-hand side of each equation equal to 0.

Stable polymorphic equilibria may exist within tumors [22, 49]. If the dynamics of the tumor can be described via equation (7) or other dynamics leading to ESSs, then these polymorphic equilibria will correspond to an ESSs [181]. Furthermore, polymorphic stability in heterogeneous tumor cell populations has been shown to exist explicitly for some cancers [17, 70].

Likely, the most influential LV competition model of cancer dynamics is that of Zhang et al. (2017) [200]. This model has been derived from the replicator dynamics in [193], while preserving their ESSs. Subsequently, the model was expanded so that it allowed for modeling abiraterone acetate treatment (further referred to as “abiraterone”), assuming that this treatment, applied together with androgen deprivation, decreased the carrying capacity of cancer cells producing testosterone. Moreover, under androgen deprivation, the carrying capacity of cancer cells dependent on testosterone was made a linear function of the density of the testosterone producing cancer cells. As such, the originally noncooperative game between the three cancer cell types includes also cooperative elements. The LV formulation has the advantage of including population dynamics providing a more realistic modeling framework. This is because treatment aims at decreasing tumor burden while keeping the proportion of treatment-resistant cancer cells low. Replicator dynamics models typically capture only the latter, unless they include birth-death processes. The LV competition model described in [200] formed the basis of the adaptive treatment protocol used in a successful clinical trial (NCT02415621) where patients received abiraterone at MTD until their initial PSA levels dropped to half and resumed only when the PSA returned to its initial value. In this way, patients had individual treatment regimens with varying length of cycles with and without treatment.

Zhang et al. simulated SoC with MTD of abiraterone combined with ADT, using clinically motivated parameters, to show how SoC strongly selects for the testosterone-independent cancer cell type, due to *competitive release* [47, 198]. This means that resistant cancer cells eventually outcompete other cells. As an alternative to the SoC, Zhang et al. proposed the above described *adaptive therapy*, whose underlying as-sumption is that in the absence of treatment resistant cells are less fit than sensitive cells (standard assumption on the fitness costs of resistance in ecology [1, 166]). This assumption can however be relaxed, as shown in [182]. Both the simulated adaptive therapy and the clinical trial treatment regiment applied arbiraterone together with ADT until the tumor volume dropped below half of its initial value, as indicated by the blood serum level of PSA. From that moment on, abiraterone was discontinued, until the tumor volume recovered to its initial level. Then the cycle was repeated. This has two anticipated effects:

1. Cancer cells are not dominated by the drug-resistant cell type.
2. The cumulative drug dose is lower.

An interesting finding is that a lower initial proportion of sensitive cells leads to longer periods of time until the PSA reaches its initial level. Adaptive therapy also results in a gradual increase of the resistant cells from cycle to cycle, but this happens much slower than with the SoC.

In summary, Zhang et al. (2017) demonstrated that this adaptive therapy regimen provides equivalent or longer time to progression (TTP) than SoC therapy under any initial conditions [200]. With their simple but effective, the adaptive therapy is not optimized; instead, the conditions to pause and restore the abiraterone treatment are rules of thumb related to the current tumor volume. The corresponding clinical trial has shown that patients’ TTP increased remarkably with this regimen. Recent updates of this clinical trial (NCT02415621) are consistent with the initial findings [200, 201]. The adaptive therapy trial prolonged TTP with less than half of the cumulative drug dose and appears to be successful for all patients that were initially responsive. Currently, the patients’ life expectancies have almost tripled. Conversely, most patients receiving the SoC have progressed.

Cunningham et al. (2018) optimized the abiraterone therapy from [200] with respect to different criteria, such as minimizing the variance of the total tumor burden [51]. This will be discussed in more detail in Section 3.

Meanwhile, West et al. (2019) investigated a *multi-drug* approach for mCRPC [190]. Clearly, this further extends the treatment options. For simplicity, they limited themselves to a two-drug approach where the secondary drug is supposed to suppress the sub-population which is resistant to the primary drug. Accordingly, they considered the treatment with docetaxel (chemotherapy) and abiraterone, considering also a cell type which is resistant to both docetaxel and abiraterone. They conducted simulations parametrized on patients that progressed in the mentioned clinical trial in Zhang et al. (NCT02415621) and reached the conclusion that the administration of docetaxel together with abiraterone would have significantly increased TTP. Based on the success of the this trial, more trials on adaptive therapies are being opened (e.g., in melanoma - NCT03543969, in thyroid cancer - NCT03630120, and also a second trial including Zhang in mCRPC - NCT03511196).

There are other examples of game-theoretic models guiding clinical trials. For example, West et al. (2019) consider a trial on stage 2/3 estrogen receptor-positive breast cancer and treatment with an aromatase inhibitor and a PD-L1 checkpoint inhibitor combination, which attempts to lower a preoperative endocrine prognostic index (PEPI) that correlates with relapse-free survival [189]. They adopted a game with a 4 × 4 fitness matrix, which was then embedded in an ecological model of tumor population-growth dynamics. The resulting model predicts evolutionary and ecological dynamics that track changes in the PEPI score. By testing out different possible treatment regimens, they proposed a therapy plan with a one-month kick start with the immune checkpoint inhibitor followed by five months of continuous combination therapy as the most effective therapy choice. Current practice either uses the drugs in combination or just uses the aromatase inhibitor.

LV models can be exploited to include also other cells interacting with the cancer cells, such as T-cells (as predators), as shown for example in [29, 145]. Alternatively, one may be interested in the role of non-immune cells, such as cancer-associated fibroblasts [191, 109] that may inhibit or facilitate the fitness of all or just some types of cancer cells. The parameters of LV models can also be inferred directly from *in vitro* experiments following a procedure similar to the game assay [138].

### 2.6. Spatial game-theoretic models and related work

There is evidence that spatial interactions among cancer cells and/or interactions of cancer cells with their environment influence *intra-tumor heterogeneity*, the spatial properties of tumors, and patient prognose [120].

In space, tumors can be viewed as complex evolving structures, consisting of cancer cells, normal cells, blood vasculature, inter-cellular spaces, and various nutrients, such as oxygen and glucose [77, 125]. Cancer cells, often of distinct types, compete for space and nutrients, and engage in direct interactions. They both contribute towards and are affected by their microenvironments, within which they consume available resources, proliferate and survive [63]. Within these neighborhoods, there are ecoevolutionary feedbacks where limiting resources impact the total abundance of cancer cells, and interactions between tumor cells influence the frequency of cell types. Moreover, spatially-explicit data, e.g., biopsies, histological samples and magnetic resonance imaging (MRI) imaging, become more and more available [158, 186] and pathologists often measure and score spatial distributions of cancer cell types, vasculature, immune cells, and other tumor properties [202, 143]. Also, cancer biologists increasingly recognize the ubiquity of spatial heterogeneity within tumors [30, 120, 165].

For these reasons, spatially-explicit models increased in popularity.

However, one has to be careful in inferring and interpreting game parameters from measurements in spatially explicit systems [105, 106].

Spatially-explicit EGT cancer models can take the form of *diffusion processes* framed as partial differential equations [175] or models can be *agent-based* [45, 117, 118]. In some special cases, it is possible to use analytic techniques to transform and solve spatially-explicit EGT models in the same way as the inviscid models we described above [110, 133, 106].

In graph-based models, the cancer cells may be represented on vertices of a network, such as Voronoi graphs [15], motivated by the claim that real biological tissues appear closest to those [50, 114]. Alternatively, individual cells may occupy a space on a spatial grid described as squares or hexagons [144, 170]. Agent-based models can also consider continuous space where the cancer cells are represented by continuously varying spatial coordinates in one, two or three dimensions, often extending the replicator dynamics (7) into spatially explicit scenarios [24, 71, 193, 194]. In this case, the interactions between the different cell types are typically more or less local and depend on how cells interact with each other, how much they can move, how local their perception of the density of the cells around them is, and/or how far from a focal cell the offspring can be placed.

For example, You et al. (2017) modelled the interaction of mCRPC cells under androgen-deprivation therapy (ADT) as an evolutionary game with three types of cancer cells (cells requiring testosterone, cells producing testosterone as public good, and cells independent of testosterone), with the fitness matrix defining cells’ probabilities of proliferating when interacting with other cells [193]. Both ESSs and properties of an agent-based continuous-space variant of this game with a birth-death process were analyzed, and their transient dynamics and eco-evolutionary equilibria were compared. The authors observed that only when interactions between cancer cells of the spatial model were global the resulting evolutionary equilibria corresponded to the ESSs of the original nonspatial game.

## 3. Game theory of cancer treatment

In case the treatment is *a priori* decided (such as in case of continuous MTD or metronomic treatment, or also with adaptive treatment with its rules decided beforehand), the physician is not a true player in the game, as they do not really optimize any objective. This was the case in the models introduced in Section 2.

Here, we consider the case where the physician becomes a true player in the game. When viewing cancer as an evolutionary game between the physician and the cancer cells, a natural question arises: Can we drive cancer into a stable state, corresponding to either a cure or a chronic disease, which is not too harmful for the patient and can be maintained at a stable tumor burden? This concept of stability corresponds to the Evolutionarily Stable Strategies introduced in Section 2. Alternatively, if cure or stable tumor burden cannot be achieved, a relevant question is whether we can maximally delay undesirable states (e.g., too high tumor burden or too high level of resistance), by more dynamical treatment protocols than currently used as SoC. To this aim, we introduce an objective function to be optimized by the physician, *Q* (**U**(·), **x**(·), **m**(·)), which varies with 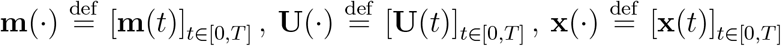. With no loss of generality, we can refer to this function as the *Quality of Life* (QoL) function of the patient. The physician’s goal is then to find the optimal **m**\* which optimizes such an objective, i.e., find

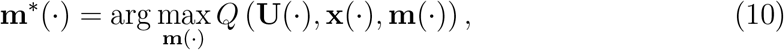

where *Q* has been decided by the physician and patient a priori. In such a situation, cancer cells are playing an evolutionary game with each other and their eco-evolutionary dynamics can still be described by equations (3) and (5). However, they become followers in a Stackelberg (i.e., leader-follower) game, with the physician as a rational leader [159]. Since the followers are evolutionary players, we call this type of games *Stackelberg evolutionary games* (SEGs), in accordance with recent research on this topic [151, 153]. It is noteworthy that the physician, as the only rational player in this SEG, can anticipate and steer the eco-evolutionary response of the cancer cells defined by (3)– (5), while the cancer cells can only adapt to the actions already taken by the physician. The theory of Stackelberg games was originally devised in economics to conceptualize interactions with an imbalance in control or power, e.g., the competition between a market leader and follower [94]. Its extension into SEGs can conceptualize not only cancer treatment, but also other problems involving a rational player interacting with an evolutionary system, e.g., pest management, fisheries management, or the control of infectious diseases [37, 38, 93, 153].

Here, we divide existing literature into two categories:

1. SEGs with cancer cells in eco-evolutionary equilibria: Here it is assumed that an equilibrium **x**\* and ESS **U**\* is reached for any given choice of **m**. Under this condition, we look for a constant **m** that maximizes *Q*(**U**\*(*m*), **x**\*(*m*), **m**).
2. SEGs where the cancer cells are assumed to be in their transient phase, with their eco-evolutionary dynamics driven by equations (3)–(5).

When it comes to the objective of the leader, we identify two important categories of literature:

1. SEGs where the leader aims at steering the cancer cells into their eco-evolutionary equilibria, assuming application of a constant dose once such an equilibrium is achieved. Here it is explicitly given up on the idea of curing and the strategy becomes “treat to contain”, similarly to what happens with chronic diseases.
2. SEGs with different objectives for the leader, such as minimization of the tumor burden, minimization of its variance, or maximization of the time to progression.

In table 1 we summarize these options and indicate the sections where each is discussed.

**Table 1:** Instances of Stackelberg evolutionary games (SEGs) of cancer treatment considered in this review

|  |  | Physician |  |
| --- | --- | --- | --- |
| | | steering to $(\mathbf{x}^*, \mathbf{U}^*)$ | another objective |
| Cancer dynamics | transient | Section 3.1 | Section 3.2 |
| | at $(\mathbf{x}^*, \mathbf{U}^*)$ | - | Section 3.3 |

### 3.1. Physician steering cancer into an ESS

Most cancer biologists and many modelers see cancer as only a transient dynamic with little focus on the idea of reaching an equilibrium (**U**\*, **x**\*) of its eco-evolutionary dynamics, and even fewer within an explicit game-theoretic setting. However, there is evidence that eco-evolutionary dynamics in cancer cells do have attractors whether reached or not [64, 80]. These works reported that if these equilibria are reached, cancer can be contained with a constant dose of treatment, lower than the MTD. [119] and [42] implied that reaching the ESSs of the cancer dynamics may be a successful strategy to keep the patient with a metastatic cancer alive. Cunningham et al. focused on steering mCRPC into its eco-evolutionary equilibria, for the model from [200], where one or several competition coefficients were increased to values above 1 [53]. This was based on a study, demonstrating that the competition coefficients among different cancer cells of different types are often above 1 [70]. Cunningham et al. first adopted a numerical *optimal control approach*, with a forward-backward sweep method to steer mCRPC to a sustainable eco-evolutionary attractor. While they showed, with perfect knowledge, that reaching such an attractor is feasible for most patients, they focused also on rules of thumb to reach these attractors, without complicated optimization of the treatment protocols [53]. They demonstrated that *dose titration*, i.e., gradual increase of treatment dose is very likely to lead to the cancer’s ESS.

### 3.2. Physician optimizing objectives other than reaching the ESS, while cancer cells are in their transient phase

Martin et al. (1992) were probably the first authors who applied optimal control in cancer treatment, with focus on various objectives for the physician [119]. They considered the population of drug-sensitive and drug-resistant cancer cells, where the goal was to slow the growth of drug-resistant cells, which also served to maximize patient survival time. Three types of tumor growth models were investigated: Gompertz, logistic, and exponential. For each model, they adopted an analytical optimal control approach to find feedback controls that specify the optimal tumor mass as a function of the size of the resistant sub-population [27, 31, 146]. With exponential and logistic tumor growth, the tumor burden during therapy had little impact on survival times. With Gompertzian tumor growth, therapies maintaining a large tumor burden doubled or even tripled patient survival time. A revolutionary finding of this paper was that maintaining a high tumor burden is optimal for Gompertz tumor growth and close to optimal for exponential and logistic tumor growth. Hence, it is not necessary to know the precise growth characteristics of a tumor to schedule anticancer drugs. Their results also implied that trying to contain the tumor may be the best strategy for keeping patients alive. A growing literature on optimal control to optimize cancer treatment has emerged as a follow-up to this work [5, 112, 134, 179].

Orlando et al. modeled the case of cancer cells trading off resistance between two different drugs with the physician solving an optimal control problem with the objective of minimizing the tumor burden [140]. They show that a relatively static treatment using both drugs at equal levels is optimal when cancer cells benefit from specializing in response to a single drug rather than a generalist resistance strategy, while a more dynamic treatment with the concentration of drugs varying over time is more effective when this multitasking is rewarding to the cancer cells [140].

Carrère focused on *in vitro* tumors, consisting of cells that were sensitive or resistant to a certain drug. The setting was similar to [119], but with parameters validated by an *in vitro* study [42]. They adopted optimal control theory and showed analytically that to reduce the tumor volume while preserving its heterogeneity, one needs to apply lower than the MTD of drugs. Warman et al. (2018) focused on a fitness matrix model of the vicious cycle of metastatic prostate cancer cells co-opting bone remodeling [187]. The authors introduced *fractionated follow up therapy* – chemotherapy where dosage is administered initially in one solid block followed by alternating smaller doses and holidays – and shiwed that it is better than either a continuous application or a periodic one. Gluzman et al. optimize treatment in a public goods model of interactions between glycolytic and acidic cells, introduced by Kaznatcheev et al. (2017) [85]. The total drug usage and time to recovery were optimized, by solving the corresponding Hamilton–Jacobi–Bellman (HJB) equation, similar to [53]. They conclude that the optimal treatment policies can significantly decrease the total amount of drugs prescribed, while also increasing the fraction of initial tumor states from which recovery is possible. This paper supports the claim that lower doses of treatment will be more effective for containing tumors than MTD.

Cunningham et al. optimize abiterone treatment from [200] using boxed-constrained optimization. They consider various objectives for the physician and show that minimization of the tumor volume variance, thus keeping the tumor burden as stable as possible, may be the best objective for keeping the patients from progressing while not applying too much drug [51].

Itik et al. introduce a model describing competition between normal cells and tumor cells. The model also includes the effects of the immune system [101]. They propose a linear time varying approximation technique to construct an optimal control strategy for the nonlinear system which is valid not only within small perturbations around the equilibrium point, but also for global dynamics of the system. The objective is to eliminate the tumor cells while minimizing the amount of drug. It should be noted, that as evolution of resistance is not included in the model, it is likely more relevant for treatment of early stage cancers, as opposed to advanced and metastatic cancers.

### 3.3. Physician optimizing various objectives, while cancer cells dynamics are at ESS

Once the ecological equilibrium **x**\* and the ESS resistance strategies **U**\* are reached, a constant dose **m**\* can keep the cancer dynamics contained [151, 152, 160]. Finding such equilibria for cancer eco-evolutionary dynamics and **m**\* for maximizing the patient’s quality of life are the main goals of [151] and [153]. For monomorphic cancer cell populations, [152] showed that less treatment leads to a higher quality of life (Fig. 1). It is to be noted that their approach considered only a monomorphic population of cancer cells, with resistance as a scalar trait. However, the fact that MTD leads to an outcome which is not better and usually much worse than the Nash equilibrium, which is in turn not better and usually much worse than the Stackelberg equilibrium, can be generalized to the situation with vector-valued traits.

**Figure 1:**
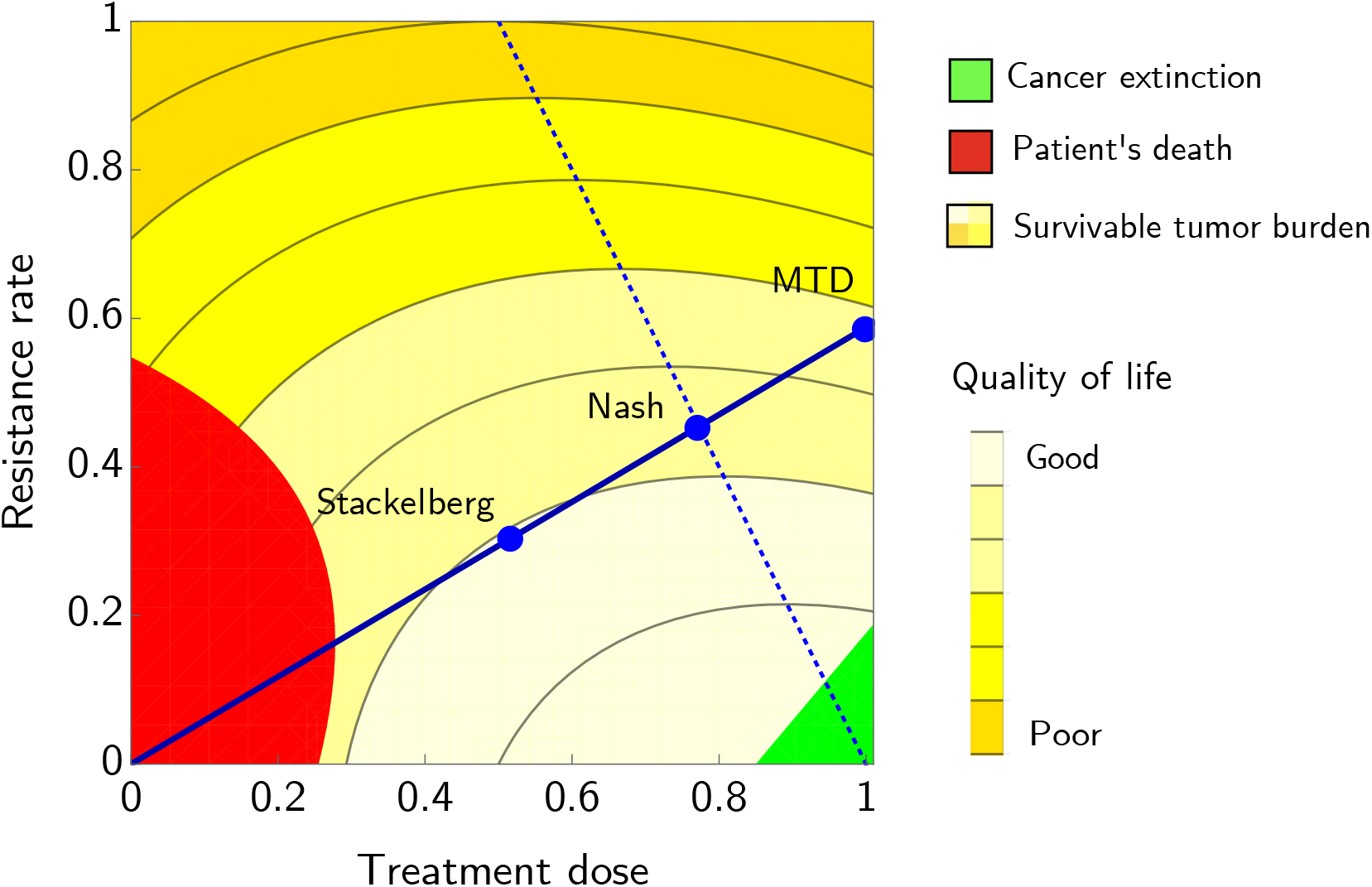
The solid line represents the best response of cancer cells to any possible treatment level *m* ∈ [0, 1], the dotted line the best response of the physician to any possible resistance level *u* ∈ [0, 1]. In the green area the population of cancer cells goes extinct, while in the red area it grows above the survival threshold of the patient and as such, it is incompatible with life. The yellow area represents the situation in-between, with different levels of quality of life. Three different equilibria of the game are presented. MTD corresponds to the case where the physician plays a fixed Maximum Tolerated Dose strategy; NASH corresponds to adjusting the dose according to the resistance rate of cancer cells, until a Nash equilibrium is reached; STACKELBERG corresponds to anticipating the followers’ resistance strategy. Adapted from [152].

## 4. Clinical relevance

Application of EGT principles in therapy, in order to anticipate and steer cancer eco-evolutionary response, is a powerful tool, but relies on our ability to estimate tumor size and composition prior to treatment. The intra-tumoral evolutionary process leads to sub-clonal diversification and generates the genetic and phenotypic intra-tumor heterogeneity, which determines the tumor composition and therefore the evolutionary state. In order to optimize the model parameters, determined by the tumor composition, monitoring of the tumor’s behavior during therapy is required. At best, this encompasses continuous surveillance of the total number of tumor cells and their cell type composition. In clinic, the personalized therapeutic strategy then needs to be optimized after every measurement, i.e., after each clinical visit. Kaznatcheev et al. [109] recently showed how to assess the game played by different cell types of non-small cell lung cancer cells *in vitro*. The game changed in response to different treatment regimens. Due to the *in vitro* set-up, the experiments could be monitored with relative ease by performing time-lapse microscopy. However, in a clinical setting the key constraint is the low amount of information available about intra-tumoral evolution and the speed of evolution during treatment. It is still challenging to identify, quantify and monitor the evolving strategy distribution in heterogeneous tumors. A sufficient technology for this is yet unavailable, however, several techniques can be proposed which we discuss in the following paragraphs.

Firstly, tissue biopsies of the primary tumor and of metastases can be sampled to reveal genetic and phenotypic differences between cancer cell types. Genetic differences are revealed by genome sequencing, while phenotypic heterogeneity is typically assessed with histology techniques and proteomics [30]. Nevertheless, to monitor the cancer cells’ response to treatment, tissues need to be isolated at the time of initial diagnosis as well as successively sampled throughout treatment. In the clinics, such repeated biopsies are not easily acceptable, due to their invasive nature and expense. Such is the case in taking biopsies of disseminated bone disease in mCRCP patients [69]. Furthermore, often only a fraction of the tumor is isolated, which does not represent the complete genomic and phenotypic landscape, and the detection of small lesions and deriving biopsies from them is a major challenge [100, 141].

Secondly, an alternative approach is based on liquid biopsies. They consist of several sources of tumor material including circulating tumor DNA (ctDNA) and circulating tumor cells (CTC). The ctDNA is a DNA released by malignant cancer cells, with diagnostic genetic and epigenetic alterations. Several studies have shown that exome-wide analysis of ctDNA may contribute to monitoring the evolution of acquired drug resistance and track the outgrowth of resistant cell types [40, 131, 132, 139]. To be able to use the genotypic information obtained from ctDNA, we need to know the relationship between mutations and their phenotypic impact, i.e., the genotype-phenotype map [4, 137]. Predicting what genotypes will eventually evolve to drive phenotypic resistance remains a significant challenge [60].

From CTCs, besides genotypic information, phenotypic information about the strategy distribution can directly be obtained for use in the EGT models. CTCs represent intact, viable non-hematological cells with malignant features. The resistant CTC populations may be phenotypically distinct from their precursors in physical size, shape and surface marker expression. For instance, Tsao et al. [178] detected tumor progression and proliferation of resistant melanoma cell types by observing surface marker up-regulation from CTCs. They saw how a widening of the signal distribution detected by spectroscopy, reflected a more heterogeneous CTC population [178]. In mCRPC, Zhang et al. [200, 201] detect testosterone producing cells by the presence of CTCs expressing CYP17A1, which is a key enzyme for androgen synthesis. Androgen receptors (AR) can also be detected and monitored in real time from mCRPC CTCs. The AR splice variant 7 was proved to be predictive of resistance to anti-AR treatment, such as ADT therapy and treatment with both abiraterone and enzalutamide [8, 127, 128, 161, 173]. Additionally, CTCs can be assayed for human epidermal growth factor receptor 2 (HER2+) in breast cancer, which contributes to treatment resistance [48, 148]. This technique may also be applied to display the strategy distribution in other cancer types, when specific up-regulation or down-regulation of specific surface markers in resistant cell types occurs.

Taking liquid biopsies and isolating CTCs, has advantages over conventional tissue biopsies since they are less invasive to the patient. Additionally, it may reflect the heterogeneity of the tumor more appropriately and it allows continuously monitoring of a patient’s tumor composition [115]. Nevertheless, the liquid biopsies provide neither spatial information nor information on the composition of individual metastatic lesions, since the primary tumor and its metastases are not measured individually. Accordingly, liquid biopsies may contain a mixture of tumor cells originating from multiple independent lesions. Analyses of primary and disseminated tumor cells show large differences in genetic variation [162], and CTCs are unlikely to represent the full spectrum of mutations and differences in protein expression in tumor lesions since CTC biopsies might only show the ‘tip of the iceberg’ [199]. While it is better to have this aggregated information as a proxy for the cancer’s evolutionary dynamics than no information at all. The information found this way can be used to measure the evolutionary states of different metastatic lesions if multiple metastatic lesions located at different sites shed CTCs homogeneously or if the variation in the composition of these lesions is low. This is e.g., shown in BRAF status concordance in primary and metastatic melanoma [33, 34] and colorectal carcinoma and KRAS mutation status in colorectal adenocarcinoma [56]. Alternatively, one needs to identify the tissue of origin of CTCs by using expression profiling of organ-specific metastatic features. Studies have shown that certain methylation patterns are tissue specific which may serve to determine the source of tumor cells or ctDNA [113, 164].

Thirdly, another approach is blood sampling to measure blood serum markers. These are biomarkers produced by specific tumor cell types. In studies by Zhang et al. [200, 201], prostate cancer volume is determined by assessing PSA levels in the blood, while in the latter research testosterone blood levels under androgen deprivation are measured. The testosterone levels are used as a proxy for the amount of testosterone-producing cancer cells. Nevertheless, whether each tumor cell produces the same amount of PSA might depend on the sensitivity of the tumor cells to androgen stimulation for the expression of PSA. Some cell types have been shown to lose sensitivity to androgen and produce even more PSA than androgen sensitive cell types [57, 102]. This feature that differs between prostate cancer cell types might provide ways to measure androgen-independent and dependent types of cancer cells. However, this is again aggregated information combining all metastatic lesions. A study in melanoma showed a higher expression of BRAF(V600E) oncoprotein in vemurafenib-resistant tumor cells compared to sensitive cells. This difference might be used for parameterizing EGT models of melanoma [169]. For other cancer types investigated in adaptive therapy studies, there is a lack of reliable biomarker presented yet.

Fourthly, modern imaging techniques are another emerging approach for gaining tumor and intra-tumoral metrics. Imaging can provide a holistic view of the entire tumor and since it is non-invasive, it is suitable for repeated monitoring. Magnetic resonance imaging (MRI) and computed tomography (CT) can be used to track spatial and temporal patterns of heterogeneity. For example, these techniques may reveal tumor habitats such as necrosis, hypoxia and vascular permeability. Such habitats may select for different cells with varying levels of responsiveness to therapy [171, 172].

Radiomics provide images of tumor habitats which seek correlations between cell phenotypes and their visual appearance. Quantitative imaging features can include shape, edge to volume ratio, texture or tissue environment. Such features can be built into predictive models relating image features to tumor cell types [3, 78]. It has already been demonstrated in studies of patients with glioblastoma multiforme that differences in cancer cell protein expression within a tumor correlates with regions of varying contrast-enhancement from MRI images [59, 96]. However, before quantitative imaging features can be used for clinical monitoring of cancer cell strategies, it needs to be ensured that specific imaging features can be linked to the underlying composition of cell types that differ in their response to treatment.

Positron Emission Tomography (PET), which can be performed along with CT or MRI scans, provides additional anatomic and spatial information. PET scans can show differential amounts and patterns of uptake of radiotracers by cells within a tumor. This might provide the ability to label and quantify the resistant as well as the sensitive cells. For example, the variability of tumor glycolytic metabolism within the same lesion can be assessed with the use of 2-flouro-2-deoxy-D-glucose F 18 ([18F]-FDG) PET imaging [28]. Uptake patterns influence patients outcome and thus provide insights into the prevalence of resistant cancer cells within the tumor. Additionally, PET imaging using fluorodihydrotestosterone F 18 ([18F]-FDHT) permits to labelling and detection of androgen receptors [196]. Accordingly, a combination of [18F]-FDG and [18F]-FDHT PET imaging can identify AR positive and negative lesions, and therefore the ability to discriminate sensitive and resistant prostate cancer cell types [68]. A radiotracer to label prostate specific membrane antibody (PSMA), a cell surface protein with high expression in prostate cancer cells, is also available for PET imaging. PSMA is expressed on nearly all prostate cancer cells, and therefore accessible to labelling [122]. Furthermore, it is under research whether the radiotracer N-succinimidyl-4-[18F]fluorobenzoate ([18F]-SFB) is suitable for labeling HER2 overexpressing cells in breast cancer [192].

Modern imaging techniques might hold more promise than tissue and liquid biopsies. This is because it can reveal relevant information about both the location of the lesions and the tumor cell types within these lesions, to reveal both tumor eco-evolutionary dynamics and spatial characteristics. Furthermore, it is non-invasive and overcomes sampling errors of biopsies. In particular, we propose PET imaging because it can provide insight in both total tumor mass and the tumor’s cell type composition. Therefore, the discovery of radiotracers, which are able to classify different tumor cell types, is of uppermost clinical importance. Nevertheless, current modern imaging studies mostly focus on how tumor metrics and not cell type composition can be used as a prognostic marker for overall survival, malignancy or therapy response. For example, Aerts et al. [2] used radiomic data from CT images of patients with early-stage NSCLC and use a response phenotype that can predict a patient’s sensitivity towards Gefitinib therapy. In order to parameterize the EGT models, all different tumor cell types in a tumor need to be identified and monitored.

It might be worthwhile to use newly developed techniques such as organoids [150] and xenografts [39] to measure cell type compositions and protein expression to monitor tumor evolution, and improve our understanding of the eco-evolutionary dynamics. Early preclinical *in vivo* studies of adaptive therapy included ovarian cancer cell line xenografts treated with carboplatin, and MDA-MB-231/luc triple-negative and MCF7 ER+ breast cancer cell lines treated with paclitaxel. In all cases adaptive therapy could stabilize tumor volume, though the underlying sub-populations were not explicitly measured [64, 80]. In both of these studies, once initial tumor volume control was achieved, it could be maintained with constant or even progressively smaller drug doses, suggestive of stable eco-evolutionary equilibria.

Once patient specific data of tumor cell types is available and monitored, it can be used to parameterize and optimize the EGT models to guide adaptive therapy protocols [95]. Subsequent measurements to inform patient specific parameters would then greatly improve modeling and predictions regarding tumor characteristics [200, 201]. After every measurement, the optimal next step in the adaptive therapeutic protocol could be calculated and used to stabilize the tumor burden, or might even be steered to create a pathway towards cure. To compare different mathematical models and seek the optimal cancer treatment, an optimal control theory approach may suffice [7, 44, 51, 54, 129, 130, 155]. Additionally, model predictive control (MPC) can use real-time monitored data to update the optimal cancer treatment. MPC involves model based control techniques which can update the model and the optimal treatment schedule with each new clinical measure [130].

Critically, a model for tumor treatment can only be as effective as its associated empirical methods allow, i.e., in order to parameterize and validate it. Data may be retrospective (histologies, radiographies, biopsies etc.) as well as derived from mouse or cell culture studies. For mapping genotypic or phenotypic data to treatment strategies, traditional statistical approaches can be used, but opportunities for machine learning / artificial intelligence are evident [103]. Furthermore, the *in vitro* and *in vivo* competition assay has been shown to be well suited to feed EGT models. Such experiments have already shown that the success of cancer lineages depends on its frequency and the frequency of all other lineages with other strategies [23, 98, 99]. General models, when augmented by case studies, will permit EGT to inform clinical practice [86].

## 5. Discussion

We have reviewed the application of EGT in modeling tumor progression with and without treatment. When considering treatment models, we made a distinction between those with *a priori* defined treatment or a null (0 dose) treatment vs. those where the physician enters the game and actively adjusts treatment strategies during the course of the treatment in response to the metrics of the cancer’s eco-evolutionary state.

We considered evolutionary approaches to treatment and anticipated increased life expectancy from evolutionary therapy as compared to the traditional therapy, when resistant types are either pre-existing or evolve in response to therapy.

The biggest obstacle to applying EGT treatment methods to clinics remains the difficulty of estimating the tumor composition which currently can be done only for some types of cancers. Therapy for such cancers is particularly suited for our approach. They have discrete cell types and can therefore be understood via simpler EGT models. We reviewed some approaches for estimating the tumor composition in Section 4.

There is a clear gap between the complexity of the models that we introduced by (3) and (5), with a resistance matrix **U**, and existing models, where either scalar or vector-valued traits are considered. We could find no research where one actually considers the resistance level of each known cancer type to each possible treatment. In all research we reviewed, resistance was either a single evolving trait of a monomorphic cancer population, a strategy within a polymorphic cancer population with one treatment, or multiple strategies of a polymorphic population with multiple treatments, where the resistance to these treatments does not evolve according to (5) but represents discrete and fixed strategies. The effects of these modeling assumptions (monomorphic or polymorphic population) and how they impact the superiority of adaptive therapy over continuous therapy with MTD have been recently investigated by Pressley et al (2021), where time to progression for monomorphic and polymorphic models was compared between adaptive therapy and MTD [147]. The most general form of the cancer model, given by (3) and (5), has recently been used by Reed et al. (2020) to model pediatric sarcomas, where tumor growth is suppressed by multiple drugs, towards which resistance is evolving (eg. vinorelbine, dactinomycin, cyclophosphamide).

Most commonly used replicator dynamics and LV equations describe only one of the equations (5) and (3). There is a rich theory for both, presumably as these models are simpler than the most general ones.

Some topics are not addressed in this paper that may become relevant for the future of game theory for cancer and its treatment. For example, we did not specify whether the cancer cells’ types correspond to genetic or non-genetic traits (e.g., epigenetics; [156]). Generally, we believe that this does not influence the conceptualization of the game-theoretic models. Furthermore, whether strategies are genetic, epigenetic or phenotipically plastic will, at times, influence evolutionary speed. It may be that the tumor micro-environment can influence the epigenetics of a cell and thereby change its type, while this would not be the case if the type is genetically determined. Future models may need to pay close attention to the role of the micro-environment on the capacity for cancer cells to switch strategies. This switching may also happen in cancer stem cells (CSCs) and have consequences for tumor heterogeneity and the composition of cancer cell types. This might be interesting to study in the context of EGT modeling and cancer therapy.

Many common cancer types are shown to be propagated by small populations of CSCs. Genetic and epigenetic alterations can lead to CSCs emerging from non-stem cells endowed with stem cell properties. Therefore, stem cell identity may not be strictly a property of that cell, but may also depend on extrinsic cues provided by the adjacent cells and microenvironment. If stemness is not an intrinsic property, the malignant cells will regenerate new cancer stem cells, even if those with stem-like properties have been eliminated. Accordingly, the stem state of a cell is a dual phenotype. Therefore, in modeling CSCs a choice must be made on whether stemness is an intrinsic property or whether cell type switching takes place or not. Analysis of the evolution of stemness can help identify whether these different types of stemness evolve according to different selective pressures, such as tissue maintenance and repair. This phenomenon also poses the question as to how non-genetically encoded plasticity will affect EGT modeling.

Close communication and collaboration between theoretical and empirical scientists will be of the utmost importance in advancing evolutionary therapies based on evolutionary game theory.

## Data Availability

There was no original data created for this manuscript.

## Acknowledgements

This research was supported by two European Union’s Horizon 2020 research and innovation programs under the Marie Skĺodowska-Curie grant agreement No’s 690817 and 955708, the Dutch National Foundation (NWO) Grants number 040.11.712, EN-WPR.020.006, OCENW.KLEIN.277, and two James S. McDonnell Foundation grants, Cancer therapy: Perturbing a complex adaptive system and a postdoctoral fellowship award (#2020-1423), a V Foundation grant, NIH/National Cancer Institute (NCI) R01CA170595, Application of Evolutionary Principles to Maintain Cancer Control (PQ21), NIH/NCI U54CA143970-05 Physical Science Oncology Network (PSON) Cancer as a complex adaptive system and Austrian Science Fund (FWF): DK W1225-B20. The last author wants to thank to her daughter Julia for keeping her awake during nights, which allowed her to work on this paper.

## Notes

### Competing Interest Statement

The authors have declared no competing interest.

### Funding Statement

This research was supported by two European Union's Horizon 2020 research and innovation programs under the Marie Sklodowska-Curie grant agreement No's 690817 and 955708, the Dutch National Foundation (NWO) Grant number OCENW.KLEIN.277, the James S. McDonnell Foundation grant, Cancer therapy: Perturbing a complex adaptive system, a V Foundation grant, NIH/National Cancer Institute (NCI) R01CA170595, Application of Evolutionary Principles to Maintain Cancer Control (PQ21), NIH/NCI U54CA143970-05 Physical Science Oncology Network (PSON) Cancer as a complex adaptive system and Austrian Science Fund (FWF): DK W1225-B20. The last author wants to thank to her daughter Julia for keeping her awake during nights, which allowed her to work on this paper.

### Author Declarations

For this paper we have not conducted any clinical trials.

## References

[1] Acar, A., Nichol, D., Fernandez-Mateos, J., Cresswell, G. D., Barozzi, I., Hong, S. P., Trahearn, N., Spiteri, I., Stubbs, M., Burke, R., et al. (2020). Exploiting evolutionary steering to induce collateral drug sensitivity in cancer. Nature Communications, 11(1):1–14.

[2] Aerts, H. J. W. L., Grossmann, P., Tan, Y., Oxnard, G. R., Rizvi, N., Schwartz, L. H., and Zhao, B. (2016). Defining a radiomic response phenotype: a pilot study using targeted therapy in NSCLC. Scientific Reports, 6:33860.

[3] Aerts, H. J. W. L., Velazquez, E. R., Leijenaar, R. T. H., Parmar, C., Grossmann, P., Carvalho, S., Bussink, J., Monshouwer, R., Haibe-Kains, B., Rietveld, D., et al. (2014). Decoding tumour phenotype by noninvasive imaging using a quantitative radiomics approach. Nature Communications, 5(1):1–9.

[4] Ahnert, S. E. (2017). Structural properties of genotype–phenotype maps. Journal of The Royal Society Interface, 14(132):20170275.

[5] Aïnseba, B. E. and Benosman, C. (2010). Optimal control for resistance and sub-optimal response in CML. Mathematical Biosciences, 227(2):81–93.

[6] Aktipis, C. A., Boddy, A. M., Jansen, G., Hibner, U., Hochberg, M. E., Maley, C. C., and Wilkinson, G. S. (2015). Cancer across the tree of life: cooperation and cheating in multicellularity. Philosophical Transactions of the Royal Society B: Biological Sciences, 370(1673):20140219.

[7] Algoul, S., Alam, M., Hossain, M., and Majumder, M. (2011). Multi-objective optimal chemotherapy control model for cancer treatment. Medical & Biological Engineering & Computing, 49:51–65.

[8] Antonarakis, E. S., Lu, C., Wang, H., Luber, B., Nakazawa, M., Roeser, J. C., Chen, Y., Mohammad, T., Chen, Y., Fedor, H. L., et al. (2014). AR-V7 and resistance to enzalutamide and abiraterone in prostate cancer. New England Journal of Medicine, 371(11):1028–1038.

[9] Antonia, S. J., Villegas, A., Daniel, D., Vicente, D., Murakami, S., Hui, R., Kurata, T., Chiappori, A., Lee, K. H., de Wit, M., Cho, B. C., Bourhaba, M., Quantin, X., Tokito, T., Mekhail, T., Planchard, D., Kim, Y.-C., Karapetis, C. S., Hiret, S., Ostoros, G., Kubota, K., Gray, J. E., Paz-Ares, L., de Castro Carpeño, J., Faivre-Finn, C., Reck, M., Vansteenkiste, J., Spigel, D. R., Wadsworth, C., Melillo, G., Taboada, M., Dennis, P. A., and Özgüroğlu, M. (2018). Overall survival with durvalumab after chemoradiotherapy in stage iii nsclc. New England Journal of Medicine, 379(24):2342–2350. PMID: 30280658.

[10] Apaloo, J. (1997). Revisiting strategic models of evolution: The concept of neighborhood invader strategies. Theoretical Population Biology, 52(1):71–77.

[11] Apaloo, J., Brown, J. S., and Vincent, T. L. (2009). Evolutionary game theory: ESS, convergence stability, and NIS. Evolutionary Ecology Research, 11(4):489–515.

[12] Archetti, M. (2013). Evolutionary game theory of growth factor production: implications for tumour heterogeneity and resistance to therapies. British Journal of Cancer, 109(4):1056–1062.

[13] Archetti, M. (2014). Evolutionary dynamics of the Warburg effect: glycolysis as a collective action problem among cancer cells. Journal of Theoretical Biology, 341:1–8.

[14] Archetti, M. (2015). Heterogeneity and proliferation of invasive cancer subclones in game theory models of the Warburg effect. Cell Proliferation, 48(2):259–269.

[15] Archetti, M. (2016). Cooperation among cancer cells as public goods games on Voronoi networks. Journal of Theoretical Biology, 396:191–203.

[16] Archetti, M. (2018). How to analyze models of nonlinear public goods. Games, 9(2):17.

[17] Archetti, M., Ferraro, D. A., and Christofori, G. (2015). Heterogeneity for IGF-II production maintained by public goods dynamics in neuroendocrine pancreatic cancer. Proceedings of the National Academy of Sciences, 112(6):1833–1838.

[18] Archetti, M. and Pienta, K. J. (2019). Cooperation among cancer cells: applying game theory to cancer. Nature Reviews Cancer, 19(2):110–117.

[19] Aupérin, A., Le Péchoux, C., Rolland, E., Curran, W. J., Furuse, K., Fournel, P., Belderbos, J., Clamon, G., Ulutin, H. C., Paulus, R., Yamanaka, T., Bozonnat, M.-C., Uitterhoeve, A., Wang, X., Stewart, L., Arriagada, R., Burdett, S., and Pignon, J.-P. (2010). Meta-analysis of concomitant versus sequential radiochemotherapy in locally advanced non–small-cell lung cancer. Journal of Clinical Oncology, 28(13):2181–2190. PMID: 20351327.

[20] Aupérin, A., Péchoux, C., Rolland, E., Curran, W. J., Furuse, K., Fournel, P., Belderbos, J., Clamon, G., Ulutin, H. C., Paulus, R., Yamanaka, T., Bozonnat, M. C., Uitterhoeve, A., Wang, X., Stewart, L., Arriagada, R., Burdett, S., and Pignon, J. P. (2010). Meta-analysis of concomitant versus sequential radiochemotherapy in locally advanced non–small-cell lung cancer. J Clin Oncol., 28(13):2181–90.

[21] Axelrod, R., Axelrod, D. E., and Pienta, K. J. (2006). Evolution of cooperation among tumor cells. Proceedings of the National Academy of Sciences, 103(36):13474–13479.

[22] Bach, L. A., Bentzen, S. M., Alsner, J., and Christiansen, F. B. (2001). An evolutionary-game model of tumour–cell interactions: possible relevance to gene therapy. European Journal of Cancer, 37(16):2116–2120.

[23] Baker, A. M., Huang, W., Wang, X. M. M., Jansen, M., Ma, X. J., Kim, J., Anderson, C. M., Wu, X., Pan, L., Su, N., Luo, Y., Domingo, E., Heide, T., Sottoriva, A., Lewis, A., Beggs, A. D., Wright, N. A., Rodriguez-Justo, M., Park, E., Tomlin-son, I., and Graham, T. A. (2017). Robust RNA-based in situ mutation detection delineates colorectal cancer subclonal evolution. Nature Communications, 8(1):1–8.

[24] Basanta, D., Hatzikirou, H., and Deutsch, A. (2008a). Studying the emergence of invasiveness in tumours using game theory. The European Physical Journal B, 63(3):393–397.

[25] Basanta, D., Scott, J. G., Fishman, M. N., Ayala, G., Hayward, S. W., and Anderson, A. R. A. (2012). Investigating prostate cancer tumour–stroma interactions: clinical and biological insights from an evolutionary game. British Journal of Cancer, 106(1):174–181.

[26] Basanta, D., Simon, M., Hatzikirou, H., and Deutsch, A. (2008b). Evolutionary game theory elucidates the role of glycolysis in glioma progression and invasion: Game theory and the role of glycolysis. Cell Proliferation, 41(6):980–987.

[27] Basar, T. and Olsder, G. J. (1999). Dynamic noncooperative game theory, volume 23. SIAM.

[28] Basu, S., Kwee, T. C., Gatenby, R. A., Saboury, B., Torigian, D. A., and Alavi, (2011). Evolving role of molecular imaging with PET in detecting and characterizing heterogeneity of cancer tissue at the primary and metastatic sites, a plausible explanation for failed attempts to cure malignant disorders. European Journal of Nuclear Medicine and Molecular Imaging, 38(6).

[29] Bayer, P., Brown, J. S., and Staňková, K. (2018). A two-phenotype model of immune evasion by cancer cells. Journal of Theoretical Biology, 455:191–204.

[30] Bedard, P. L., Hansen, A. R., Ratain, M. J., and Siu, L. L. (2013). Tumour heterogeneity in the clinic. Nature, 501(7467):355–364.

[31] Bellman, R. (1957). Dynamic Programming. Princeton University Press, Princeton, New Jersey.

[32] Bhattacharya, R., Vander Velde, R., Marusyk, V., Desai, B., Kaznatcheev, A., Marusyk, A., and Basanta, D. (2020). Understanding the evolutionary games in nsclc microenvironment. bioRxiv.

[33] Bledsoe, J. R., Kamionek, M., and Mino-Kenudson, M. (2014). BRAF V600E immunohistochemistry is reliable in primary and metastatic colorectal carcinoma regardless of treatment status and shows high intratumoral homogeneity. The American Journal of Surgical Pathology, 38(10):1418.

[34] Boursault, L., Haddad, V., Vergier, B., Cappellen, D., Verdon, S., Bellocq, J., Jouary, T., and Merlio, J. (2013). Tumor homogeneity between primary and metastatic sites for BRAF status in metastatic melanoma determined by immuno-histochemical and molecular testing. PLOS One, 8(8):e70826.

[35] Broom, M. and Rychtář, J. (2013). Game-theoretical models in biology. Chapman & Hall/CRC Mathematical and Computational Biology series. CRC Press, Taylor and Francis Group, Boca Raton, FL.

[36] Brown, J. S. (2016). Why Darwin would have loved evolutionary game theory. Proceedings of the Royal Society B, 283(1838):20160847.

[37] Brown, J. S. and Parman, A. O. (1993). Consequences of size-selective harvesting as an evolutionary game. In The exploitation of evolving resources, pages 248–261. Springer.

[38] Brown, J. S. and Staňková, K. (2017). Game theory as a conceptual framework for managing insect pests. Current Opinion in Insect Science, 21:26–32.

[39] Bruna, A., Rueda, O. M., Greenwood, W., Batra, A. S., Callari, M., Batra, R. N., Pogrebniak, K., Sandoval, J., Cassidy, J. W., Tufegdzic-Vidakovic, A., et al. (2016). A biobank of breast cancer explants with preserved intra-tumor heterogeneity to screen anticancer compounds. Cell, 167(1):260–274.

[40] Burger, J. A., Landau, D. A., Taylor-Weiner, A., Bozic, I., Zhang, H., Sarosiek, K., Wang, L., Stewart, C., Fan, J., Hoellenriegel, J., et al. (2016). Clonal evolution in patients with chronic lymphocytic leukaemia developing resistance to BTK inhibition. Nature Communications, 7(1):1–13.

[41] Campbell, B. B., Light, N., Fabrizio, D., Zatzman, M., Fuligni, F., de Borja, R., Davidson, S., Edwards, M., Elvin, J. A., Hodel, K. P., Zahurancik, W. J., Suo, Z., Lipman, T., Wimmer, K., Kratz, C. P., Bowers, D. C., Laetsch, T. W., Dunn, G. P., Johanns, T. M., Grimmer, M. R., Smirnov, I. V., Larouche, V., Samuel, D., Bronsema, A., Osborn, M., Stearns, D., Raman, P., Cole, K. A., Storm, P. B., Yalon, M., Opocher, E., Mason, G., Thomas, G. A., Sabel, M., George, B., Ziegler, D. S., Lindhorst, S., Issai, V. M., Constantini, S., Toledano, H., Elhasid, R., Farah, R., Dvir, R., Dirks, P., Huang, A., Galati, M. A., Chung, J., Ramaswamy, V., Irwin, M. S., Aronson, M., Durno, C., Taylor, M. D., Rechavi, G., Maris, J. M., Bouffet, E., Hawkins, C., Costello, J. F., Meyn, M. S., Pursell, Z. F., Malkin, D., Tabori, U., and Shlien, A. (2017). Comprehensive Analysis of Hypermutation in Human Cancer. Cell, 171(5):1042–1056.

[42] Carrére, C. (2017). Optimization of an in vitro chemotherapy to avoid resistant tumours. Journal of Theoretical Biology, 413:24–33.

[43] Chang, C. H., Qiu, J., O’Sullivan, D., Buck, M. D., Noguchi, T., Curtis, J. D., Chen, Q., Gindin, M., Gubin, M. M., van der Windt, G. J. W., Tonc, E., Schreiber, R. D., Pearce, E. J., and Pearce, E. L. (2015). Metabolic Competition in the Tumor Microenvironment Is a Driver of Cancer Progression. Cell, 162(6):1229–1241.

[44] Chen, T., Kirkby, N. F., and Jena, R. (2012). Optimal dosing of cancer chemotherapy using model predictive control and moving horizon state/parameter estimation. Computer Methods and Programs in Biomedicine, 108(3):973–983.

[45] Chen, Y., Wang, H., Zhang, J., Chen, K., and Li, Y. (2015). Simulation of avascular tumor growth by agent-based game model involving phenotype-phenotype interactions. Scientific Reports, 5:17992.

[46] Conitzer, V. (2013). The exact computational complexity of evolutionarily stable strategies. In International Conference on Web and Internet Economics, pages 96–108. Springer.

[47] Connell, J. (1961). The influence of interspecific competition and other factors on the distribution of the barnacle Chthamalus stellatus. Ecology, 42(4):710–723.

[48] Creighton, C. J., Massarweh, S., Huang, S., Tsimelzon, A., Hilsenbeck, S. G., Osborne, C. K., Shou, J., Malorni, L., and Schiff, R. (2008). Development of resistance to targeted therapies transforms the clinically associated molecular profile subtype of breast tumor xenografts. Cancer Research, 68(18):7493–7501.

[49] Cross, W. C. H., Graham, T. A., and Wright, N. A. (2016). New paradigms in clonal evolution: punctuated equilibrium in cancer. The Journal of Pathology, 240(2):126–136.

[50] Csikász-Nagy, A., Escuderoand, L. M., and Guillaud, M. e. a. (2013). Cooperation and competition in the dynamics of tissue architecture during homeostasis and tumorigenesis. Seminars in Cancer Biology, 23:293–298.

[51] Cunningham, J. J., Brown, J. S., Gatenby, R. A., and Staňková, K. (2018). Optimal control to develop therapeutic strategies for metastatic castrate resistant prostate cancer. Journal of Theoretical Biology, 459:67–78.

[52] Cunningham, J. J., Gatenby, R. A., and Brown, J. S. (2011). Evolutionary Dynamics in Cancer Therapy. Molecular Pharmaceutics, 8(6):2094–2100.

[53] Cunningham, J. J., Thuijsman, F., Peeters, R., Viossat, Y., Brown, J. S., Gatenby, R. A., and Staňková, K. (2020). Optimal Control to Reach Eco-Evolutionary Stability in Metastatic Castrate Resistant Prostate Cancer. PLOS One, 15(12).

[54] Czakó, B., Sápi, J., and Kovács, L. (2017). Model-based optimal control method for cancer treatment using model predictive control and robust fixed point method. In 2017 IEEE 21st International Conference on Intelligent Engineering Systems (INES), pages 000271–000276. IEEE.

[55] Darwin, C. (1859). On the Origin of Species by Means of Natural Selection.

[56] Murray. de Macedo, M. P., Melo, F. M., Ribeiro, H. S. C., Marques, M. C., Kagohara, L. T., Begnami, M. D., Neto, J. C., Ribeiro, J. S., Soares, F. A., Carraro, D. M., et al. (2017). KRAS mutation status is highly homogeneous between areas of the primary tumor and the corresponding metastasis of colorectal adenocarcinomas: one less problem in patient care. American Journal of Cancer Research, 7(9):1978.

[57] Denmeade, S. R., Sokoll, L. J., Dalrymple, S., Rosen, D. M., Gady, A. M., Bruzek, D., Ricklis, R. M., and Isaacs, J. T. (2003). Dissociation between androgen responsiveness for malignant growth vs. expression of prostate specific differentiation markers PSA, hK2, and PSMA in human prostate cancer models. The Prostate, 54(4):249–257.

[58] Dieckmann, U. and Law, R. (1996). The dynamical theory of coevolution: a derivation from stochastic ecological processes. Journal of Mathematical Biology, 34(5-6):579–612.

[59] Diehn, M., Nardini, C., Wang, D. S., McGovern, S., Jayaraman, M., Liang, Y., Aldape, K., Cha, S., and Kuo, M. D. (2008). Identification of noninvasive imaging surrogates for brain tumor gene-expression modules. Proceedings of the National Academy of Sciences, 105(13):5213–5218.

[60] Dienstmann, R., Rodon, J., Barretina, J., and Tabernero, J. (2013). Genomic medicine frontier in human solid tumors: prospects and challenges. Journal of Clinical Oncology, 31(15):1874–1884.

[61] Dingli, D., Chalub, F. A. C. C., Santos, F. C., Van Segbroeck, S., and Pacheco, J. M. (2009). Cancer phenotype as the outcome of an evolutionary game between normal and malignant cells. British Journal of Cancer, 101(7):1130–1136.

[62] Dujon, A. M., Aktipis, A., Alix-Panabiéres, C., Amend, S. R., Boddy, A. M., Brown, J. S., Capp, J.-P., DeGregori, J., Ewald, P., Gatenby, R., Gerlinger, M., Giraudeau, M., Hamede, R. K., Hansen, E., Kareva, I., Maley, C. C., Marusyk, A., McGranahan, N., Metzger, M. J., Nedelcu, A. M., Noble, R., Nunney, L., Pienta, K. J., Polyak, K., Pujol, P., Read, A. F., Roche, B., Sebens, S., Solary, E., Staňková, K., Swain Ewald, H., Thomas, F., and Ujvari, B. (2020). Identifying key questions in the ecology and evolution of cancer. Evolutionary Applications, 0000:1–16.

[63] Egeblad, M., Nakasone, E. S., and Werb, Z. (2010). Tumors as organs: complex tissues that interface with the entire organism. Developmental Cell, 18(6):884–901.

[64] Enriquez-Navas, P. M., Kam, Y., Das, T., Hassan, S., Silva, A., Foroutan, P., Ruiz, E., Martinez, G., Minton, S., Gillies, R., and Gatenby, R. A. (2016). Exploiting evolutionary principles to prolong tumor control in preclinical models of breast cancer. Science Translational Medicine, 8(327):327ra24.

[65] Farrokhian, N., Maltas, J., Ellsworth, P., Durmaz, A., Dinh, M., Hitomi, M., Kaznatcheev, A., Marusyk, A., and Scott, J. G. (2020). Dose dependent evolutionary game dynamics modulate competitive release in cancer therapy. bioRxiv. DOI 10.1101/2020.09.18.303966.

[66] Fisher, R. A. (1930). The Genetical Theory of Natural Selection. Clarendon Press, Oxford, UK.

[67] Fortunato, A., Boddy, A. M., Mallo, D., Aktipis, C. A., Maley, C. C., and Pepper, J. W. (2017). Natural Selection in Cancer Biology: From Molecular Snowflakes to Trait Hallmarks. Cold Spring Harbor Perspectives in Medicine, 7(2):a029652.

[68] Fox, J. J., Gavane, S. C., Blanc-Autran, E., Nehmeh, S., Gönen, M., Beattie, B., Vargas, H. A., Schöder, H., Humm, J. L., Fine, S. W., et al. (2018). Positron emission tomography/computed tomography–based assessments of androgen receptor expression and glycolytic activity as a prognostic biomarker for metastatic castrationresistant prostate cancer. JAMA Oncology, 4(2):217–224.

[69] Fraser-Hill, M. A. and Renfrew, D. L. (1992). Percutaneous needle biopsy of musculoskeletal lesions. 1. Effective accuracy and diagnostic utility. AJR. American Journal of Roentgenology, 158(4):809–812.

[70] Freischel, A. R., Damaghi, M., Cunningham, J. J., Ibrahim-Hashim, A., Gillies, R. J., Gatenby, R. A., and Brown, J. S. (2020). Frequency-dependent interactions determine outcome of competition between two breast cancer cell lines. bioRxiv. DOI 10.1101/2020.03.06.979518.

[71] Gallaher, J. and Anderson, A. R. A. (2013). Evolution of intratumoral phenotypic heterogeneity: the role of trait inheritance. Interface Focus, 3(4):20130016.

[72] Gatenby, R. (1995). Models of Tumor-Host Interaction as Competing Populations: Implications for Tumor Biology and Treatment. Journal of Theoretical Biology, 176(4):447–455.

[73] Gatenby, R. (2009a). A change of strategy in the war on cancer. Nature, 459:508–509.

[74] Gatenby, R. A. (2009b). A change of strategy in the war on cancer. Nature, 459(7246):508–509.

[75] Gatenby, R. A., Artzy-Randrup, Y., Epstein, T., Reed, D. R., and Brown, J. S. (2020). Eradicating metastatic cancer and the eco-evolutionary dynamics of anthropocene extinctions. Cancer Research, 80(3):613–623.

[76] Gatenby, R. A. and Brown, J. S. (2018). The evolution and ecology of resistance in cancer therapy. Cold Spring Harbor Perspectives in Medicine, 8(3):a033415.

[77] Gatenby, R. A., Cunningham, J. J., and Brown, J. S. (2014). Evolutionary triage governs fitness in driver and passenger mutations and suggests targeting never mutations. Nature Communications, 5:1–9.

[78] Gatenby, R. A., Grove, O., and Gillies, R. J. (2013). Quantitative imaging in cancer evolution and ecology. Radiology, 269(1):8–14.

[79] Gatenby, R. A. and Maini, P. K. (2003). Mathematical oncology: cancer summed up. Nature, 421(6921):321–321.

[80] Gatenby, R. A., Silva, A. S., Gillies, R. J., and Frieden, B. R. (2009). Adaptive therapy. Cancer Research, 69(11):4894–4903.

[81] Gatenby, R. A. and Vincent, T. L. (2003). An evolutionary model of carcinogenesis. Cancer Research, 63(19):6212–6220.

[82] Gatenby, R. A., Zhang, J., and Brown, J. S. (2019). First Strike–Second Strike Strategies in Metastatic Cancer: Lessons from the Evolutionary Dynamics of Extinction. Cancer Research, 79(13):3174–3177.

[83] Geritz, S. A. H., Meszéna, G., Metz, J. A. J., et al. (1998). Evolutionarily singular strategies and the adaptive growth and branching of the evolutionary tree. Evolutionary Ecology, 12(1):35–57.

[84] Gerlee, P. and Altrock, P. M. (2017). Extinction rates in tumour public goods games. Journal of The Royal Society Interface, 14(134):20170342.

[85] Gluzman, M., Scott, J. G., and Vladimirsky, A. (2020). Optimizing adaptive cancer therapy: dynamic programming and evolutionary game theory. Proceedings of the Royal Society B, 287(1925):20192454.

[86] Grunspan, D. Z., Nesse, R. M., Barnes, M. E., and Brownell, S. E. (2018). Core principles of evolutionary medicine: A Delphi study. Evolution, Medicine and Public Health, 1:13–23.

[87] Halloway, A., Staňková, K., and Brown, J. S. (2019). Non-Equilibrial Dynamics in Under-Saturated Communities. Technical report, Evolutionary Biology. DOI 10.1101/834838.

[88] Hamilton, W. D. (1963). The evolution of altruistic behavior. American Naturalist, 97(896):354–356.

[89] Hamilton, W. D. (1967). Extraordinary Sex Ratios. Science, 156(3774):477–488.

[90] Hanahan, D. and Weinberg, R. A. (2000). The Hallmarks of Cancer. Cell, 100(1):57–70.

[91] Hanahan, D. and Weinberg, R. A. (2011). Hallmarks of Cancer: The Next Generation. Cell, 144(5):646–674.

[92] Hastings, A. and Gross, L. (2012). Encyclopedia of Theoretical Ecology. Number 4 in Encyclopedias of the Natural World. University of California Press.

[93] Heino, M. (1998). Management of evolving fish stocks. Canadian Journal of Fisheries and Aquatic Sciences, 55(8):1971–1982.

[94] Hicks, J. R. and von Stackelberg, H. (1935). Marktform und Gleichgewicht. The Economic Journal, 45(178):334.

[95] Ho, D. (2020). Artificial intelligence in cancer therapy. Science, 367(6481):982–983.

[96] Hobbs, S. K., Shi, G., Homer, R., Harsh, G., Atlas, S. W., and Bednarski, M. D. (2003). Magnetic resonance image-guided proteomics of human glioblastoma multiforme. Journal of Magnetic Resonance Imaging, 18(5):530–536.

[97] Hofbauer, J. and Sigmund, K. (1998). Evolutionary Games and Population Dynamics. Cambridge University Press.

[98] Huang, W., Haubold, B., Hauert, C., and Traulsen, A. (2012). Emergence of stable polymorphisms driven by evolutionary games between mutants. Nature Communications, 3(1):1–7.

[99] Huang, W., Traulsen, A., Werner, B., Hiltunen, T., and Becks, L. (2017). Dynamical trade-offs arise from antagonistic coevolution and decrease intraspecific diversity. Nature Communications, 8(1):1–8.

[100] Ilié, M. and Hofman, P. (2016). Pros: Can tissue biopsy be replaced by liquid biopsy? Translational Lung Cancer Research, 5(4):420.

[101] Itik, M., Salamci, M., and Banks, S. (2009). Optimal control of drug therapy in cancer treatment. Nonlinear Analysis: Theory, Methods & Applications, 71(12):e1473 –e1486.

[102] Jia, L. and Coetzee, G. A. (2005). Androgen Receptor–Dependent PSA Expression in Androgen-Independent Prostate Cancer Cells Does Not Involve Androgen Receptor Occupancy of the PSA Locus. Cancer Research, 65(17):8003–8008.

[103] Jochems, A., Deist, T. M., van Soest, J., Eble, M., Bulens, P., Coucke, P., Dries, W., Lambin, P., and Dekker, A. (2016). Distributed learning: Developing a predictive model based on data from multiple hospitals without data leaving the hospital - A real life proof of concept. Radiotherapy and Oncology: Journal of the European Society for Therapeutic Radiology and Oncology, 121(3):459–467.

[104] Kaznatcheev, A. (2016). Lotka-volterra, replicator dynamics, and stag hunting bacteria. Theory, Evolution, and Games Group.

[105] Kaznatcheev, A. (2017). Two conceptions of evolutionary games: reductive vs effective. bioRxiv, page 231993.

[106] Kaznatcheev, A. (2018). Effective games and the confusion over spatial structure. Proceedings of the National Academy of Sciences, 115(8):E1709–E1709.

[107] Kaznatcheev, A. (2019). Computational complexity as an ultimate constraint on evolution. Genetics, 212(1):245–265.

[108] Kaznatcheev, A. (2020). Evolution is exponentially more powerful with frequency-dependent selection. bioRxiv.

[109] Kaznatcheev, A., Peacock, J., Basanta, D., Marusyk, A., and Scott, J. G. (2019). Fibroblasts and alectinib switch the evolutionary games played by non-small cell lung cancer. Nature Ecology & Evolution, 3:450–456.

[110] Kaznatcheev, A., Scott, J. G., and Basanta, D. (2015). Edge effects in game-theoretic dynamics of spatially structured tumours. Journal of The Royal Society Interface, 12(108):20150154.

[111] Kaznatcheev, A., Vander Velde, R., Scott, J. G., and Basanta, D. (2017). Cancer treatment scheduling and dynamic heterogeneity in social dilemmas of tumour acidity and vasculature. British Journal of Cancer, 116:785–792.

[112] Ledzewicz, U. and Schaettler, H. (2016). Optimizing Chemotherapeutic Anti-cancer Treatment and the Tumor Microenvironment: An Analysis of Mathematical Models. Advances in Experimental Medicine and Biology, 936:209–223.

[113] Lehmann-Werman, R., Neiman, D., Zemmour, H., Moss, J., Magenheim, J., Vaknin-Dembinsky, A., Rubertsson, S., Nellgård, B., Blennow, K., Zetterberg, H., et al. (2016). Identification of tissue-specific cell death using methylation patterns of circulating DNA. Proceedings of the National Academy of Sciences, 113(13):E1826–E1834.

[114] Lewis, F. T. (1928). The correlation between cell division and the shapes and sizes of prismatic cells in the epidermis of Cucumis. The Anatomical Record, 38:341–376.

[115] Lianidou, E. S., Strati, A., and Markou, A. (2014). Circulating tumor cells as promising novel biomarkers in solid cancers. Critical Reviews in Clinical Laboratory Sciences, 51(3):160–171.

[116] Lotka, A. J. (1926). Elements of physical biology. Science Progress in the Twentieth Century (1919–1933), 21(82):341–343.

[117] Macklin, P. and Edgerton, M. E. (2010). Agent-based Cell Modeling: Application to Breast Cancer. Cambridge University Press.

[118] Mansury, Y., Diggory, M., and Deisboeck, T. S. (2006). Evolutionary game theory in an agent-based brain tumor model: exploring the ‘genotype–phenotype’link. Journal of Theoretical Biology, 238(1):146–156.

[119] Martin, R. B., Fisher, M. E., Minchin, R. F., and Teo, K. L. (1992). Optimal control of tumor size used to maximize survival time when cells are resistant to chemotherapy. Mathematical Biosciences, 110(2):201–219.

[120] Marusyk, A., Almendro, V., and Polyak, K. (2012). Intra-tumour heterogeneity: a looking glass for cancer? Nature Reviews Cancer, 12(5):323–334.

[121] Marusyk, A., Tabassum, D. P., Altrock, P. M., Almendro, V., Michor, F., and Polyak, K. (2014). Non-cell autonomous tumor-growth driving supports sub-clonal heterogeneity. Nature, 514:54–58.

[122] Maurer, T., Eiber, M., Schwaiger, M., and Gschwend, J. E. (2016). Current use of PSMA–PET in prostate cancer management. Nature Reviews Urology, 13(4):226–235.

[123] Maynard Smith, J. (1982). Evolution and the Theory of Games. Cambridge University Press.

[124] Maynard Smith, J. and Price, G. R. (1973). The Logic of Animal Conflict. Nature, 246(5427):15–18.

[125] Merlo, L., Pepper, J., Reid, B., and Maley, C. (2006). Cancer as an evolutionary and ecological process. Nature Reviews Cancer, 6(12):924–935.

[126] Metz, J. A. J., Geritz, S. A. H., Meszéna, G., Jacobs, F. J. A., and Van Heer-waarden, J. S. (1995). Adaptive dynamics: a geometrical study of the consequences of nearly faithful reproduction. Stochastic and Spatial Structures of Dynamical Systems, Proceedings of the Royal Dutch Academy of Science (KNAW Verhandelingen), North Holland, Amsterdam, pages 183–231.

[127] Miyamoto, D. T., Lee, R. J., Stott, S. L., Ting, D. T., Wittner, B. S., Ulman, M., Smas, M. E., Lord, J. B., Brannigan, B. W., Trautwein, J., et al. (2012). Androgen receptor signaling in circulating tumor cells as a marker of hormonally responsive prostate cancer. Cancer Discovery, 2(11):995–1003.

[128] Miyamoto, D. T., Zheng, Y., Wittner, B. S., Lee, R. J., Zhu, H., Broderick, K. T., Desai, R., Fox, D. B., Brannigan, B. W., Trautwein, J., et al. (2015). RNA-Seq of single prostate CTCs implicates noncanonical Wnt signaling in antiandrogen resistance. Science, 349(6254):1351–1356.

[129] Moradi, H., Vossoughi, G., and Salarieh, H. (2013). Optimal robust control of drug delivery in cancer chemotherapy: a comparison between three control approaches. Computer Methods and Programs in Biomedicine, 112(1):69–83.

[130] Muros, F. M., Maestre, J. M., You, L., and Staňková, K. (2017). Model predictive control for optimal treatment in a spatial cancer game. In 2017 IEEE 56th Annual Conference on Decision and Control (CDC), pages 5539–5544.

[131] Murtaza, M., Dawson, S., Pogrebniak, K., Rueda, O. M., Provenzano, E., Grant, J., Chin, S., Tsui, D. W. Y., Marass, F., Gale, D., et al. (2015). Multifocal clonal evolution characterized using circulating tumour DNA in a case of metastatic breast cancer. Nature Communications, 6(1):1–6.

[132] Murtaza, M., Dawson, S., Tsui, D. W. Y., Gale, D., Forshew, T., Piskorz, A. M., Parkinson, C., Chin, S., Kingsbury, Z., Wong, A. S. C., et al. (2013). Non-invasive analysis of acquired resistance to cancer therapy by sequencing of plasma DNA. Nature, 497(7447):108–112.

[133] Nanda, M. and Durrett, R. (2017). Spatial evolutionary games with weak selection. Proceedings of the National Academy of Sciences, 114(23):6046–6051.

[134] Nanda, S., Moore, H., and Lenhart, S. (2007). Optimal control of treatment in a mathematical model of chronic myelogenous leukemia. Mathematical Biosciences, 210(1):143–156.

[135] Nash, J. F. (1950). Equilibrium points in N-person games. Proceedings of the National Academy of Sciences, 36:48–49.

[136] Nesse, R. M., Bergstrom, C. T., Ellison, P. T., Flier, J. S., Gluckman, P., Govindaraju, D. R., Niethammer, D., Omenn, G. S., Perlman, R. L., Schwartz, M. D., Thomas, M. G., Stearns, S. C., and Valle, D. (2010). Making evolutionary biology a basic science for medicine. Proceedings of the National Academy of Sciences, 107(Suppl 1):1800–1807.

[137] Nichol, D., Robertson-Tessi, M., Anderson, A. R. A., and Jeavons, P. (2019). Model genotype–phenotype mappings and the algorithmic structure of evolution. Journal of the Royal Society Interface, 16(160):20190332.

[138] Noble, R. J., Walther, V., Roumestand, C., Hibner, U., Hochberg, M. E., and Lassus, P. (2020). Paracrine behaviors arbitrate parasite-like interactions between tumor subclones. bioRxiv.

[139] Ojamies, P. N., Kontro, M., Edgren, H., Ellonen, P., Lagström, S., Almusa, H., Miettinen, T., Eldfors, S., Tamborero, D., Wennerberg, K., et al. (2017). Monitoring therapy responses at the leukemic subclone level by ultra-deep amplicon resequencing in acute myeloid leukemia. Leukemia, 31(5):1048–1058.

[140] Orlando, P. A., Gatenby, R. A., and Brown, J. S. (2012). Cancer treatment as a game: integrating evolutionary game theory into the optimal control of chemotherapy. Physical Biology, 9(6):065007.

[141] Overman, M. J., Modak, J., Kopetz, S., Murthy, R., Yao, J. C., Hicks, M. E., Abbruzzese, J. L., and Tam, A. L. (2013). Use of research biopsies in clinical trials: are risks and benefits adequately discussed? Journal of Clinical Oncology, 31(1):17.

[142] Oxnard, G. R. (2016). The cellular origins of drug resistance in cancer. Nature medicine, 22(3):232.

[143] Patel, A. P., Tirosh, I., Trombetta, J. J., Shalek, A. K., Gillespie, S. M., Wakimoto, H., Cahill, D. P., Nahed, B. V., Curry, W. T., and Martuza, R. L. (2014). Single-cell RNA-seq highlights intratumoral heterogeneity in primary glioblastoma. Science, 344(6190):1396–1401.

[144] Perfahl, H., Byrne, H. M., Chen, T., Estrella, V., Alarcón, T., Lapin, A., Gatenby, R. A., Gillies, R. J., Lloyd, M. C., and Maini, P. K. (2011). Multiscale modelling of vascular tumour growth in 3D: the roles of domain size and boundary conditions. PLOS One, 6(4):e14790.

[145] Pilon-Thomas, S., Kodumudi, K. N., El-Kenawi, A. E., Russell, S., Weber, A. M., Luddy, K., Damaghi, M., Wojtkowiak, J. W., Mulé, J. J., Ibrahim-Hashim, A., and Gillies, R. J. (2016). Neutralization of Tumor Acidity Improves Antitumor Responses to Immunotherapy. Cancer Research, 76(6):1381–1390.

[146] Pontryagin, L. S., Boltianski, V. G., Gamkrelidze, R. V., Mishchenko, E. F., and Brown, D. E. (1964). The mathematical theory of optimal processes. A Pergamon Press book.

[147] Pressley, M., Salvioli, M., Lewis, D. B., Richards, C. L., Brown, J. S., and Stankova, K. (2021). Speed of evolutionary dynamics of drug resistance in cancer cells impacts superiority of adaptive therapy over maximum tolerable dose. Under review.

[148] Punnoose, E. A., Atwal, S. K., Spoerke, J. M., Savage, H., Pandita, A., Yeh, R., Pirzkall, A., Fine, B. M., Amler, L. C., Chen, D. S., et al. (2010). Molecular biomarker analyses using circulating tumor cells. PLOS One, 5(9):e12517.

[149] Rockne, R. C., Hawkins-Daarud, A., Swanson, K. R., Sluka, J. P., Glazier, J. A., Macklin, P., Hormuth II, D. A., Jarrett, A. M., Lima, E. A., Oden, J. T., et al. (2019). The 2019 mathematical oncology roadmap. Physical biology, 16(4):041005.

[150] Sachs, N., de Ligt, J., Kopper, O., Gogola, E., Bounova, G., Weeber, F., Balgobind, A. V., Wind, K., Gracanin, A., Begthel, H., et al. (2018). A living biobank of breast cancer organoids captures disease heterogeneity. Cell, 172(1-2):373–386.

[151] Salvioli, M. (2020). Game theory for improving medical decisions and managing biological systems. PhD thesis, Politecnico di Milano, Milano, Italy.

[152] Salvioli, M., Brown, J. S., Dubbeldam, J. L. A., and Staňková, K. (2020). Evolutionary treatment of metastatic cancer. Under review.

[153] Salvioli, M., Dubbeldam, J. L. A., Staňková, K., and Brown, J. S. (2021). Fisheries management as a Stackelberg Evolutionary Game: Finding an evolutionarily enlightened strategy. Plos One, 16(1):e0245255.

[154] Sartakhti, J. S., Manshaei, M. H., and Archetti, M. (2018). Game theory of tumor–stroma interactions in multiple myeloma: effect of nonlinear benefits. Games, 9(2):32.

[155] Sharifi, N., Ozgoli, S., and Ramezani, A. (2017). Multiple model predictive control for optimal drug administration of mixed immunotherapy and chemotherapy of tumours. Computer Methods and Programs in Biomedicine, 144:13–19.

[156] Sharma, S., Kelly, T. K., and Jones, P. A. (2010). Epigenetics in cancer. Carcinogenesis, 31(1):27–36.

[157] Sinervo, B. and Lively, C. M. (1996). The rock–paper–scissors game and the evolution of alternative male strategies. Nature, 380(6571):240–243.

[158] Sottoriva, A., Spiteri, I., Piccirillo, S. G. M., Touloumis, A., Collins, V. P., Marioni, J. C., Curtis, C., Watts, C., and Tavaré, S. (2013). Intratumor heterogeneity in human glioblastoma reflects cancer evolutionary dynamics. Proceedings of the National Academy of Sciences, 110(10):4009–4014.

[159] Staňková, K. (2019). Resistance games. Nature Ecology & Evolution, 3(3):336–337.

[160] Staňková, K., Brown, J. S., Dalton, W. D., and Gatenby, R. A. (2019). Optimizing Cancer Treatment Using Game Theory. JAMA Oncology, 5(1):96–103.

[161] Steinestel, J., Luedeke, M., Arndt, A., Schnoeller, T. J., Lennerz, J. K., Wurm, C., Maier, C., Cronauer, M. V., Steinestel, K., and Schrader, A. J. (2019). Detecting predictive androgen receptor modifications in circulating prostate cancer cells. Oncotarget, 10(41):4213.

[162] Stoecklein, N. H. and Klein, C. A. (2010). Genetic disparity between primary tumours, disseminated tumour cells, and manifest metastasis. International Journal of Cancer, 126(3):589–598.

[163] Sun, C., Wang, L., Huang, S., Heynen, G. J. J. E., Prahallad, A., Robert, C., Haanen, J., Blank, C., Wesseling, J., Willems, S. M., Zecchin, D., Hobor, S., Bajpe, P. K., Lieftink, C., Mateus, C., Vagner, S., Grernrum, W., Hofland, I., Schlicker, A., Wessels, L. F. A., Beijersbergen, R. L., Bardelli, A., Di Nicolantonio, F., Eggermont, A. M. M., and Bernards, R. (2014). Reversible and adaptive resistance to BRAFV600E inhibition in melanoma. Nature, 508(7494):118–122.

[164] Sun, K., Jiang, P., Chan, K. C. A., Wong, J., Cheng, Y. K. Y., Liang, R. H. S., Chan, W., Ma, E. S. K., Chan, S. L., Cheng, S. H., et al. (2015). Plasma DNA tissue mapping by genome-wide methylation sequencing for noninvasive prenatal, cancer, and transplantation assessments. Proceedings of the National Academy of Sciences, 112(40):E5503–E5512.

[165] Swanton, C. (2012). Intratumor heterogeneity: evolution through space and time. Cancer Research, 72(19):4875–4882.

[166] Szakács, G., Hall, M. D., Gottesman, M. M., Boumendjel, A., Kachadourian, R., Day, B. J., Baubichon-Cortay, H., and Di Pietro, A. (2014). Targeting the Achilles Heel of Multidrug-Resistant Cancer by Exploiting the Fitness Cost of Resistance. Chemical Reviews, 114(11):5753–5774.

[167] Taylor, C., Fudenberg, D., Sasaki, A., and Nowak, M. A. (2004). Evolutionary Game Dynamics in Finite Populations. Bulletin of Mathematical Biology, 66(6):1621–1644.

[168] Taylor, P. D. and Jonker, L. B. (1978). Evolutionary stable strategies and game dynamics. Mathematical Biosciences, 40(1–2):145–156.

[169] Thakur, M. D., Salangsang, F., Landman, A. S., Sellers, W., Pryer, N. K., Levesque, M. P., Dummer, R., McMahon, M., and Stuart, D. D. (2013). Modelling vemurafenib resistance in melanoma reveals a strategy to forestall drug resistance. Nature, 494(7436):251–255.

[170] Thalhauser, C. J., Lowengrub, J. S., Stupack, D., and Komarova, N. L. (2010). Selection in spatial stochastic models of cancer: migration as a key modulator of fitness. Biology Direct, 5(1):11–21.

[171] Thews, O., Nowak, M., Sauvant, C., and Gekle, M. (2011). Hypoxia-induced extracellular acidosis increases p-glycoprotein activity and chemoresistance in tumors in vivo via p38 signaling pathway. In Oxygen Transport to Tissue XXXII, pages 115–122. Springer.

[172] Tixier, F., Le Rest, C. C., Hatt, M., Albarghach, N., Pradier, O., Metges, J., Corcos, L., and Visvikis, D. (2011). Intratumor heterogeneity characterized by textural features on baseline 18F-FDG PET images predicts response to concomitant radiochemotherapy in esophageal cancer. Journal of Nuclear Medicine, 52(3):369–378.

[173] Todenhöfer, T., Azad, A., Stewart, C., Gao, J., Eigl, B. J., Black, P. C., Joshua, M., and Chi, K. N. (2016). Correlation of a novel whole blood RT-PCR assay measuring AR-V7 expression with outcomes in metastatic castration-resistant prostate cancer (mCRPC) patients treated with abiraterone acetate (ABI).

[174] Tollis, M., Boddy, A. M., and Maley, C. C. (2017). Peto’s Paradox: how has evolution solved the problem of cancer prevention? BMC Biology, 15(1):1–5.

[175] Tomasetti, C., Vogelstein, B., and Parmigiani, G. (2013). Half or more of the somatic mutations in cancers of self-renewing tissues originate prior to tumor initiation. Proceedings of the National Academy of Sciences, 110(6):1999–2004.

[176] Tomlinson, I. P. M. (1997). Game-theory models of interactions between tumour cells. European Journal of Cancer, 33(9):1495–1500.

[177] Trivers, R. L. (1971). The evolution of reciprocal altruism. Quarterly Review of Biology, 46(1):35–57.

[178] Tsao, S. C., Wang, J., Wang, Y., Behren, A., Cebon, J., and Trau, M. (2018). Characterising the phenotypic evolution of circulating tumour cells during treatment. Nature Communications, 9(1):1–10.

[179] Villasana, M., Ochoa, G., and Aguilar, S. (2010). Modeling and optimization of combined cytostatic and cytotoxic cancer chemotherapy. Artificial Intelligence in Medicine, 50(3):163–173.

[180] Vincent, T. L. and Brown, J. S. (2005). Evolutionary game theory, natural selection, and Darwinian dynamics. Cambridge University Press.

[181] Viossat, Y. (2015). Evolutionary dynamics and dominated strategies. Economic Theory Bulletin, 3:91–113.

[182] Viossat, Y. and Noble, R. J. (2020). The logic of containing tumors. bioRxiv. DOI 10.1101/2020.01.22.915355.

[183] Volterra, V. (1927). Variazioni e fluttuazioni del numero d’individui in specie animali conviventi. Accademia dei Lincei.

[184] von Neumann, J. (1928). Zur Theorie der Gesellschaftsspiele. Mathematische Annalen, 100(1):295–320.

[185] von Neumann, J. and Morgenstern, O. (1944). Theory of Games and Economic Behavior. Princeton University Press.

[186] Waclaw, B., Bozic, I., Pittman, M. E., Hruban, R. H., Vogelstein, B., and Nowak, M. A. (2015). A spatial model predicts that dispersal and cell turnover limit intratumour heterogeneity. Nature, 525(7568):261–264.

[187] Warman, P. I., Kaznatcheev, A., Araujo, A., Lynch, C. C., and Basanta, D. (2018). Fractionated follow-up chemotherapy delays the onset of resistance in bone metastatic prostate cancer. Games, 9(2):19.

[188] West, J., Ma, Y., and Newton, P. K. (2018). Capitalizing on competition: An evolutionary model of competitive release in metastatic castration resistant prostate cancer treatment. Journal of Theoretical Biology, 455:249–260.

[189] West, J., Robertson-Tessi, M., Luddy, K., Park, D. S. Williamson, D. F. K., Harmon, C., Khong, H. T. S. B. J., and Anderson, A. R. A. (2019a). The Immune Checkpoint Kick Start: Optimization of Neoadjuvant Combination Therapy Using Game Theory. Clinical Cancer Informatics, 3:1–12. doi: 10.1200/CCI.18.00078.

[190] West, J. B., Dinh, M. N., Brown, J. S., Zhang, J., Anderson, A. R. A., and Gatenby, R. A. (2019b). Multidrug Cancer Therapy in Metastatic Castrate-Resistant Prostate Cancer: An Evolution-Based Strategy. Clinical Cancer Research, 25(14):4413–4421.

[191] Wu, A., Liao, D., Tlsty, T. D., Sturm, J. C., and Austin, R. H. (2014). Game theory in the death galaxy: interaction of cancer and stromal cells in tumour microenvironment. Interface Focus, 4(4):20140028.

[192] Xavier, C., Blykers, A., Vaneycken, I., D’Huyvetter, M., Heemskerk, J., Lahoutte, T., Devoogdt, N., and Caveliers, V. (2016). 18F-nanobody for PET imaging of HER2 overexpressing tumors. Nuclear Medicine and Biology, 43(4):247–252.

[193] You, L., Brown, J. S., Thuijsman, F., Cunningham, J. J., Gatenby, R. A., Zhang, J., and Staňková, K. (2017). Spatial vs. non-spatial eco-evolutionary dynamics in a tumor growth model. Journal of Theoretical Biology, 435:78–97. http://dx.doi.org/10.1016/j.jtbi.2017.08.022.

[194] You, L., von Knobloch, M., Lopez, T., Peschen, V., Radcliffe, S., Koshy Sam, P., Thuijsman, F., Stankova, K., and Brown, J. S. (2019). Including Blood Vasculature into a Game-Theoretic Model of Cancer Dynamics. Games, 10(1).

[195] Zahavi, A. (1975). Mate selection - a selection for a handicap. Journal of Theoretical Biology, 53(1):205–214.

[196] Zanzonico, P. B., Finn, R., Pentlow, K. S., Erdi, Y., Beattie, B., Akhurst, T., Squire, O., Morris, M., Scher, H., McCarthy, T., et al. (2004). PET-based radiation dosimetry in man of 18F-fluorodihydrotestosterone, a new radiotracer for imaging prostate cancer. Journal of Nuclear Medicine, 45(11):1966–1971.

[197] Zeeman, E. C. (1980). Population dynamics from game theory. In Global theory of dynamical systems, pages 471–497. Springer.

[198] Zeilinger, A. R., Olson, D. M., and Andow, D. A. (2016). Competitive release and outbreaks of non-target pests associated with transgenic Bt cotton. Ecological Applications, 26:1047–1054.

[199] Zhang, C., Guan, Y., Sun, Y., Ai, D., and Guo, Q. (2016). Tumor heterogeneity and circulating tumor cells. Cancer Letters, 374(2):216–223.

[200] Zhang, J., Cunningham, J. J., Brown, J. S., and Gatenby, R. A. (2017). Integrating evolutionary dynamics into treatment of metastatic castrate-resistant prostate cancer. Nature Communications, 8(1):1–9.

[201] Zhang, J., Fishman, M. N., Brown, J. S., and Gatenby, R. A. (2019). Integrating evolutionary dynamics into treatment of metastatic castrate-resistant prostate cancer (mCRPC): Updated analysis of the adaptive abiraterone (abi) study (NCT02415621). Journal of Clinical Oncology, 37(15):5041–5041.

[202] Zhang, J., Fujimoto, J., Zhang, J., Wedge, D. C., Song, X., Zhang, J., Seth, S., Chow, C., Cao, Y., and Gumbs, C. (2014). Intratumor heterogeneity in localized lung adenocarcinomas delineated by multiregion sequencing. Science, 346(6206):256–259.

